# Genome-wide association study of REM sleep behavior disorder in Parkinson’s disease

**DOI:** 10.1101/2023.05.24.23289628

**Authors:** Yuri L. Sosero, Karl Heilbron, Pierre Fontanillas, Lucy Norcliffe-Kaufmann, Eric Yu, Uladzislau Rudakou, Jennifer A. Ruskey, Kathryn Freeman, Farnaz Asayesh, Kajsa Brolin, Maria Swanberg, Huw R Morris, Lesley Wu, Raquel Real, Lasse Pihlstrøm, Manuela Tan, Thomas Gasser, Kathrin Brockmann, Hui Liu, Michele T.M. Hu, Donald G. Grosset, Simon J.G. Lewis, John B. Kwok, Pau Pastor, Ignacio Alvarez, Matej Skorvanek, Alexandra Lackova, Miriam Ostrozovicova, Mie Rizig, 23andMe Research Team, The International Parkinson’s Disease Genomics Consortium, Lynne Krohn, Ziv Gan-Or

## Abstract

**Objective:** REM sleep behavior disorder (RBD) is a prodromal synucleinopathy, reported in a subset of Parkinson’s disease (PD) patients, and associated with neuropsychiatric symptoms in PD. We aimed to compare the genetic background of PD patients with probable RBD (PD+RBD) and PD patients without probable RBD (PD-RBD). Furthermore, we examined genetic correlations and potential causal associations between multiple neuropsychiatric traits and PD+RBD.

**Methods:** We performed a genome-wide association study (GWAS) including 5,403 PD+RBD and 13,020 PD-RBD. To test for genetic correlations and potential causal associations between neuropsychiatric traits and PD+RBD, we used linkage disequilibrium score regression and Mendelian randomization.

**Results:** The *SNCA* locus was associated with PD+RBD compared to PD-RBD (rs10005233, OR=1.21, 95% CI=1.16-1.27, *p*=1.81e-15). Further examination of known genetic loci associated with PD from the most recent PD GWAS in Europeans and Asians identified additional variants associated with reduced risk for PD+RBD: two in the *SNCA* locus (rs5019538-G, OR=0.85, 95% CI=0.81-0.89, *p*=2.46E-10; rs356182-G, OR=0.89, 95% CI=0.84-0.95, *p*=0.0001), and one in the *LRRK2* locus (rs34637584, p.G2019S, OR=0.41, 95% CI=0.28-0.61, *p*=1.04E-5). We found a potential genetic correlation between attention deficit hyperactivity disorder (ADHD) and PD+RBD, which was not statistically significant after correction for multiple comparisons. No causative association emerged between PD and neuropsychiatric traits.

**Interpretation:** Genetic variants contribute to the occurrence of RBD in PD, further distinguishing between the PD+RBD and PD-RBD subtypes. Understanding the mechanisms underlying these genetic associations could contribute to the development of subtype-specific treatments.

## INTRODUCTION

Rapid-eye-movement (REM) sleep behavior disorder (RBD) is a parasomnia characterized by the absence of muscle atonia during REM sleep and dreams enactment.^1^ When no neurological conditions or other concomitant factors are identified, it is referred to as isolated/idiopathic RBD (iRBD). ^2^ iRBD is typically considered a prodromal stage of synucleinopathies, as about 80%-90% of the cases convert to either Parkinson’s disease (PD), dementia with Lewy bodies (DLB) or, more rarely, multiple system atrophy (MSA).^3, 4^ These disorders are all characterized by the accumulation of alpha-synuclein, encoded by the *SNCA* gene.^5^ RBD is therefore a key prodromal clinical marker of synucleinopathies, and its presence is also associated with a distinctive, more severe clinical presentation. In PD patients with RBD (approximately 25%-58% of cases^6^), RBD is associated with a more malignant phenotype, characterized by faster progression^7^ and greater frequency and/or severity of neuropsychiatric manifestations, including cognitive decline, hallucinations, depression, anxiety and apathy.^8–11^

In recent years, it was shown that the genetic background of iRBD only partially overlaps with that of PD or DLB. Genes such as *GBA1*,^12^ *TMEM175*^13^ and *SNCA*^14^ are important across all conditions,^15, 16^ whereas other genes including *LRRK2*,^17^ *APOE*^18^ and familial PD genes,^19^ seem to not have a major role in iRBD. A recent RBD genome-wide association study (GWAS) identified 5 risk loci associated with RBD,^20^ namely *GBA1, TMEM175, INPPSF, SNCA* and *SCARB2*. Notably, the variants associated with RBD in the *SNCA* and *SCARB2* regions were different and independent to those associated with PD,^15, 20^ supporting RBD as a distinctive subtype, with specific genetic and clinical correlates.

In the current study, we aimed to examine whether there are differences in the genetics of PD with probable RBD (PD+RBD) compared to PD without RBD (PD-RBD). For this purpose, we performed a GWAS including 18,423 patients, composed of PD+RBD patients (N=5,403), and PD-RBD patients (N=13,020). To further explore the relationships between RBD and neuropsychiatric manifestations in PD, we performed genetic correlation and Mendelian randomization (MR) analyses using the GWAS summary statistics of the current study and multiple neuropsychiatric conditions.

## METHODS

### Population

The study population included 18,423 PD patients (detailed in Table 1), of whom 5,403 had probable RBD (PD+RBD) and were treated as cases, whereas 13,020 did not (PD-RBD) and were treated as controls. Probable RBD was defined using either the RBD single-question screen (RBD1Q)^21^ or the RBD screening questionnaire (RBDSQ),^22^ both of which show high sensitivity and specificity in PD patients.^23^ We refer to iRBD when RBD occurs prior to the neurodegeneration and to RBD for subjects with RBD regardless of the time of onset of neurodegeneration. PD was diagnosed by movement disorder specialists according to the UK Brain Bank^24^ or International Parkinson Disease and Movement Disorders Society criteria.^25^ The 23andMe cohort had self-reported a diagnosis of PD as well as RBD and/or dream re-enactment behaviors.

**Table 1.**
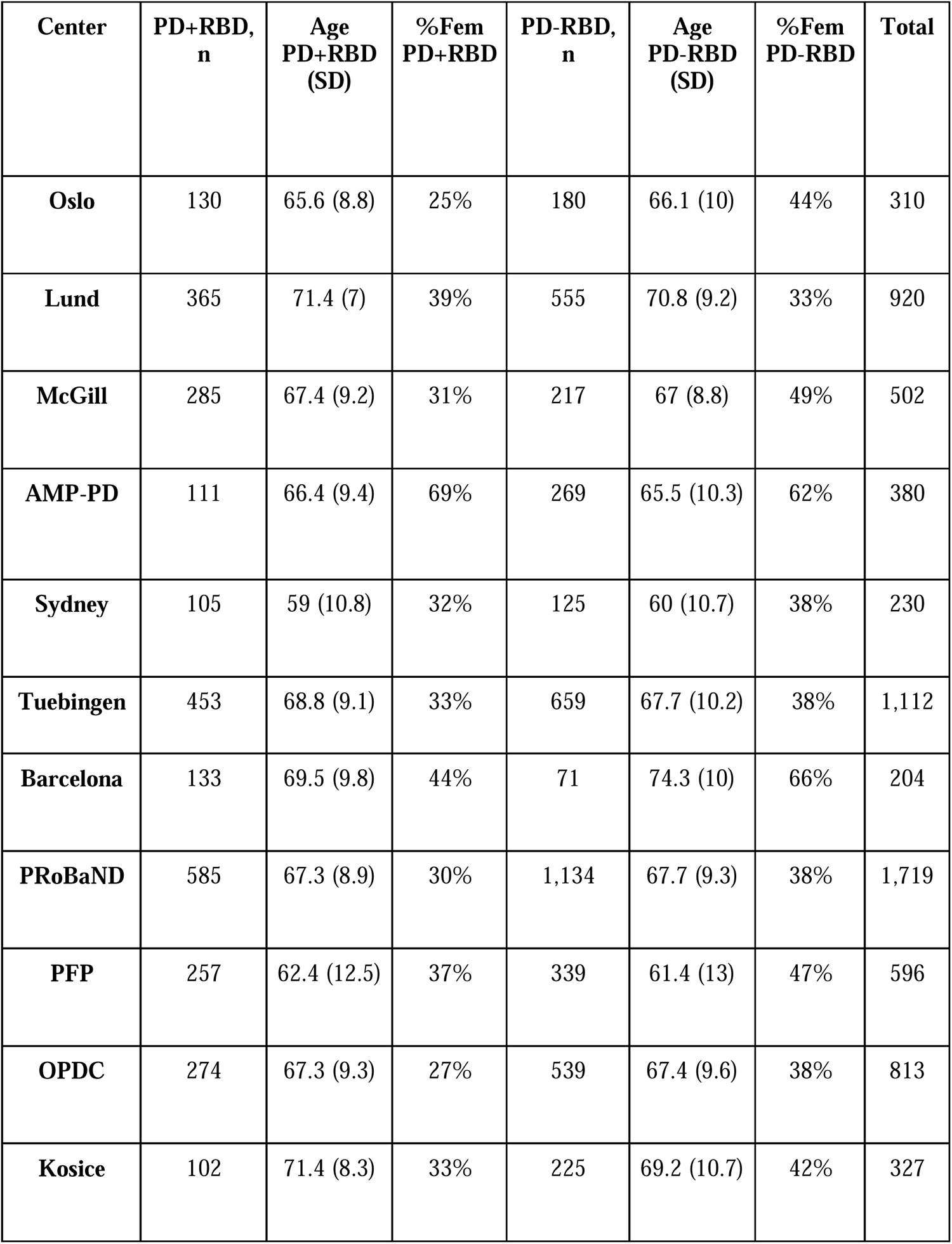
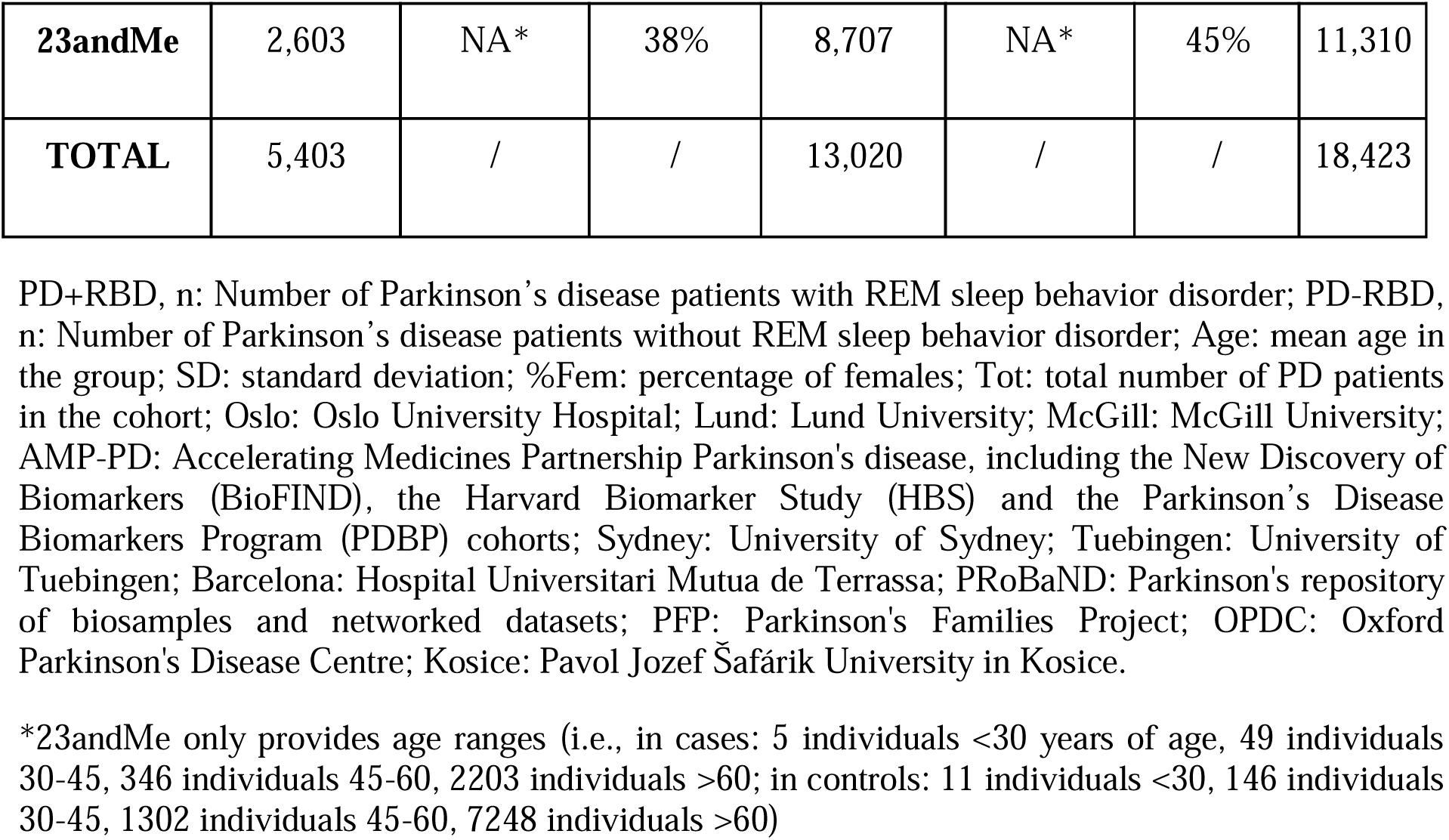
Demographic characteristics of PD patients in the individual cohorts.

The participants were of European ancestry and their clinical and genetic data were collected from 15 different cohorts (Table 1), 11 of which are from the International Parkinson’s Disease Genomics Consortium (IPDGC), three cohorts are from the Accelerating Medicines Partnership Parkinson’s disease (AMP-PD, https://amp-pd.org/) and one cohort was collected and analyzed by 23andMe Inc. (https://www.23andme.com/research/). The Central European Group on Genetics of Movement Disorders (CEGEMOD) contributed to the Kosice cohort.

### Genetic analysis

#### 23andMe

Participants provided informed consent and volunteered to participate in the research online, under a protocol approved by the external AAHRPP-accredited IRB, Ethical & Independent (E&I) Review Services. As of 2022, E&I Review Services is part of Salus IRB (https://www.versiticlinicaltrials.org/salusirb). DNA extraction and genotyping were performed on saliva samples by National Genetics Institute (NGI), a CLIA-licensed clinical laboratory and a subsidiary of Laboratory Corporation of America. Samples were genotyped on one of five genotyping platforms. The v1 and v2 platforms were variants of the Illumina HumanHap550 + BeadChip, including about 25,000 custom single nucleotide polymorphisms (SNPs) selected by 23andMe, with a total of about 560,000 SNPs. The v3 platform was based on the Illumina OmniExpress+ BeadChip, with custom content to improve the overlap with the 23andMe v2 array, with a total of about 950,000 SNPs. The v4 platform was a fully customized array, including a lower redundancy subset of v2 and v3 SNPs with additional coverage of lower-frequency coding variation, and about 570,000 SNPs. The v5 platform, in current use, is an Illumina Infinium Global Screening Array (∼640,000 SNPs) supplemented with ∼50,000 SNPs of custom content. This array was specifically designed to better capture global genetic diversity and to help standardize the platform for genetic research. Samples that failed to reach 98.5% call rate were re-analyzed. Individuals whose analyses failed repeatedly were re-contacted by 23andMe customer service to provide additional samples.

Participants were restricted to European ancestry through an analysis of local ancestry.^26^ A support vector machine (SVM) to classify individual haplotypes into one of 31 reference populations was used (https://www.23andme.com/ancestry-composition-guide/). The SVM classifications are then fed into a hidden Markov model (HMM) that accounts for switch errors and incorrect assignments, and gives probabilities for each reference population in each window. Finally, we used simulated admixed individuals to recalibrate the HMM probabilities so that the reported assignments are consistent with the simulated admixture proportions. A maximal set of unrelated individuals was chosen for each analysis using a segmental identity-by-descent (IBD) estimation algorithm.^27^

We phased participant data using either an internally-developed tool, Finch (V1-V4 genotyping arrays) or Eagle2 (V5 genotyping array).^28^ Finch implements the Beagle haplotype graph-based phasing algorithm, modified to separate the haplotype graph construction and phasing steps.^29^ It extends the Beagle model to accommodate genotyping error and recombination, to handle cases where there are no consistent paths through the haplotype graph for the individual being phased. We constructed haplotype graphs for European and non-European samples on each 23andMe genotyping platform from a representative sample of genotyped individuals, and then performed out-of-sample phasing of all genotyped individuals against the appropriate graph. For the X-chromosome, we built separate haplotype graphs for the non-pseudoautosomal region and each pseudoautosomal region, and these regions were phased separately.

Imputation panels created by combining multiple smaller panels have been shown to give better imputation performance than the individual constituent panels alone.^30^ To that end, we combined the May 2015 release of the 1000 Genomes Phase 3 haplotypes with the UK10K imputation reference panel to create a single unified imputation reference panel.^31, 32^ Multiallelic sites with N alternate alleles were split into N separate biallelic sites. We then removed any site whose minor allele appeared in only one sample. For each chromosome, we used Minimac3 to impute the reference panels against each other, reporting the best-guess genotype at each site.^33^ This gave us calls for all samples over a single unified set of variants. We then joined these together to get, for each chromosome, a single VCF with phased calls at every site for 6,285 samples.

In preparation for imputation, we split each chromosome of the reference panel into chunks of no more than 300,000 variants, with overlaps of 10,000 variants on each side. We used a single batch of 10,000 individuals to estimate Minimac3 imputation model parameters for each chunk.^33^ We imputed phased participant data against the chunked merged reference panel using Minimac3, treating males as homozygous pseudo-diploids for the non-pseudoautosomal region. Throughout, we treated structural variants and small indels the same as SNPs.

We excluded SNPs that: 1) had a call rate < 90%, 2) had a Hardy-Weinberg p < 10–20 in people with predominantly European ancestry, 3) were only genotyped on the V1 and/or V2 platforms, 4) were found on the mitochondrial chromosome or the Y-chromosome, 5) failed a test for parent-offspring transmission (specifically, we regressed the child’s allele count against the mean parental allele count and excluded SNPs with fitted < 0.6 and p < 10–20 for a test of < 1), 6) had an association with genotype date (p < 10–50 by ANOVA of SNP genotypes against a factor dividing genotyping date into 20 roughly equal-sized buckets), 7) had a large sex effect (ANOVA of SNP genotypes, r2 > 0.1), or 8) had probes matching multiple genomic positions in the reference genome.

We excluded SNPs with imputed r2 < 0.3, as well as SNPs that had strong evidence of a platform batch effect. For each SNP we identified the largest sub-set of the data passing other quality control criteria based on their original genotyping platform – either v2+v3+v4+v5, v4+v5, v4, or v5 only – and computed association test results for the largest passing set. The batch effect test is an F test from an ANOVA of the SNP dosages against a factor representing the V4 or V5 platform; we excluded results with p < 10–50.

Across both genotyped and imputed GWAS results, we excluded SNPs that had sample size of less than 20% of the total GWAS sample size. We also removed SNPs that did not converge during logistic regression, as identified by abs (effect)>10 or stderr > 10 on the log-odds scale. If SNPs were both genotyped and imputed, and they passed QC for both, we used results from the imputed analysis. After quality control, we had analysed 904,040 genotyped SNPs and 25,208,208 imputed SNPs.

GWAS was performed using logistic regression adjusted for age, sex, top five principal components as well as the genotype platform to account for genotype batch effects. The significance threshold was set at *p*<5×10E-8.

#### Other cohorts

Genotyping in the different centers was performed using the OmniExpress, NeuroX or Global Screening (GSA) GWAS array according to the manufacturer’s instructions (Illumina Inc.). Parkinson’s Families Project (PFP) was genotyped with NeuroChip, Parkinson’s repository of biosamples and networked datasets (PRoBaND) with HumanCoreExome array, and Oxford Parkinson’s Disease Centre (OPDC) with either HumanCoreExome-12 v.1.1 or Infinium HumanCoreExome-24v.1.1 arrays. Quality control was performed following standard pipelines (detailed in https://github.com/neurogenetics/GWAS-pipeline) using plink 1.9.^34^ In brief, we filtered out heterozygosity outliers using an F-statistic cut-off of <-0.15 or >0.15. Individuals with a variant call rate <95% and sex mismatch were excluded. Variants missing in >5% of the participants, with disparate missingness between cases and controls (*p*<1E-04), or significantly deviating from the Hardy-Weinberg equilibrium in controls (p<1E-04) were also removed. We used GCTA to check for relatedness closer than first cousins between participants (pihat>0.125). We performed imputation using the Michigan imputation server (https://imputationserver.sph.umich.edu/index.html#) with the Haplotype Reference Consortium reference panel r1.1 2016 under default settings. Ancestry outliers were detected using HapMap3 principal component analysis (PCA) data in R version 4.0.1. After imputation, we selected variants with R^2^>0.8 and a minor allele frequency (MAF)>0.01, while retaining variants that have strong pathogenic implications in PD (i.e., the LRRK2 p.G2019S variant and the GBA1 p.N370S, p.E326K and p.T369M variants).

To test for genetic associations to RBD in PD, we performed GWAS using logistic regression comparing PD+RBD and PD-RBD adjusted for age at RBD questionnaire administration, sex and principal components. The significance threshold was set at *p*<5×10E-8. The analyses were performed separately in each cohort and the results were then meta-analyzed with a fixed-effect model using METAL (https://genome.sph.umich.edu/wiki/METAL_Documentation).^35^ To identify any possible secondary associations hidden by the principal signals of the GWAS, we also performed Conditional and Joint - Genome-wide Complex Trait Analysis (COJO-GCTA), a method that harnesses a conditional stepwise regression approach to identify independent associations (https://yanglab.westlake.edu.cn/software/gcta/#Overview).^36^

### Genetic correlation

To investigate the potential genetic correlation between the presence of RBD in PD and known neuropsychiatric conditions we used linkage-disequilibrium score regression (LDSC) on LDHub (http://ldsc.broadinstitute.org/ldhub/).^37^ The neuropsychiatric traits we analyzed include epilepsy, headache, amyotrophic lateral sclerosis, cognitive decline, Alzheimer’s disease, Parkinson’s disease, dementia with Lewy bodies, alcohol dependence, cannabis dependence, attention deficit hyperactivity disorder, Tourette syndrome, anorexia nervosa, post-traumatic syndrome, schizophrenia, bipolar disorder, obsessive-compulsive disorder, autism spectrum disorder and major depressive disorder. Summary statistics for the compared traits were accessed through the LDHub platform or downloaded from publicly available sources, then formatted and analyzed using LDHub python v2.7 scripts (https://github.com/bulik/ldsc/wiki/). Positive correlation indicates association with PD+RBD, and negative correlation indicates association with PD-RBD.

### Mendelian randomization

To assess any possible causal association between neuropsychiatric disorders and the presence of RBD in PD we performed Mendelian randomization (MR).^38^ In brief, this method harnesses summary statistics from an exposure (the neuropsychiatric traits, in this case) and an outcome (the presence of RBD in PD) and uses the statistically significant variants from the former as instrumental variables (IVs) to infer a potential causative association with the latter. This approach mimics randomized control trials, since genetics is randomly assigned at conception and unaffected by the environment.^39^ The neuropsychiatric traits for this analysis were selected based on relevance to RBD comorbidities, known neuropsychiatric manifestations in PD or with clinical relevance to PD. They include Alzheimer’s disease, dementia with Lewy Bodies, schizophrenia, major depressive disorder and bipolar disorder. We used the TwoSampleMR R package (https://mrcieu.github.io/TwoSampleMR/)^40^ to perform MR analyses, including sensitivity analyses, tests assessing pleiotropy and heterogeneity between IVs, in R version 4.0.1 according to protocols previously established.^41^ Sensitivity analyses included MR Egger, inverse variance weighted (IVW), weighted median, simple mode and weighted mode. Steiger filtering was also performed to check for reverse causality. Summary statistics were downloaded by the MRBase GWAS catalog (http://www.mrbase.org/) and the Psychiatric Genomics Consortium (https://pgc.unc.edu/) publicly available database. To calculate the power to detect an odds ratio=1.2 we used an online Mendelian Randomization power calculation tool (https://sb452.shinyapps.io/power/).^42^

## RESULTS

### Genome-wide association study identifies the *SNCA* and *LRRK2* loci as modifiers of risk for RBD in PD

To assess whether genetics can affect the risk of RBD in PD we performed GWAS between PD+RBD (N=5,403) and PD-RBD (N=13,020). We evaluated the genomic inflation using quantile-quantile plots (Q-Q plots) and the lambda factor, showing no inflation (lambda=0.994, lambda1000=0.999), (Supplementary Fig 1).

We found that rs10005233, in the 5’ region of the *SNCA* locus, was associated with PD+RBD (OR=1.21, 95% CI=1.16-1.27, *p*=1.81E-15, Fig 1). No secondary signal was detected in the GCTA-COJO analysis at a GWAS significance level. We also examined the 92 variants associated with PD in the most recently published GWAS in Europeans^15^ and Asians^43^ (Table 2, Supplementary Table 1). Using Bonferroni correction based on the number of these variants (α/number of variants=0.00054), we identified three associations. Two were variants in the *SNCA* locus, independent of each other, whose minor alleles were associated with decreased risk for PD+RBD (rs5019538-G, OR=0.85, 95% CI=0.81-0.89, *p*=2.46E-10 and rs356182-G, OR=0.89, 95% CI=0.84-0.95, *p*=0.0001), and one was the LRRK2 p.G2019S variant, also associated with a reduced risk for PD+RBD (rs34637584, OR=0.41, 95% CI=0.28-0.61, *p*=1.04E-5, the carrier frequency for this variant across the different cohorts is detailed in Table 3). These three variants were associated with increased risk for PD in the most recent GWAS.^15, 43^ *GBA1* variants did not show significant associations with PD+RBD (Supplementary Table 2). Additional potential associations in the *SETD1A*, *SPPL2B*, *CRHR1* and *LINC00693* loci should be further studied (Table 2).

**Fig 1.**
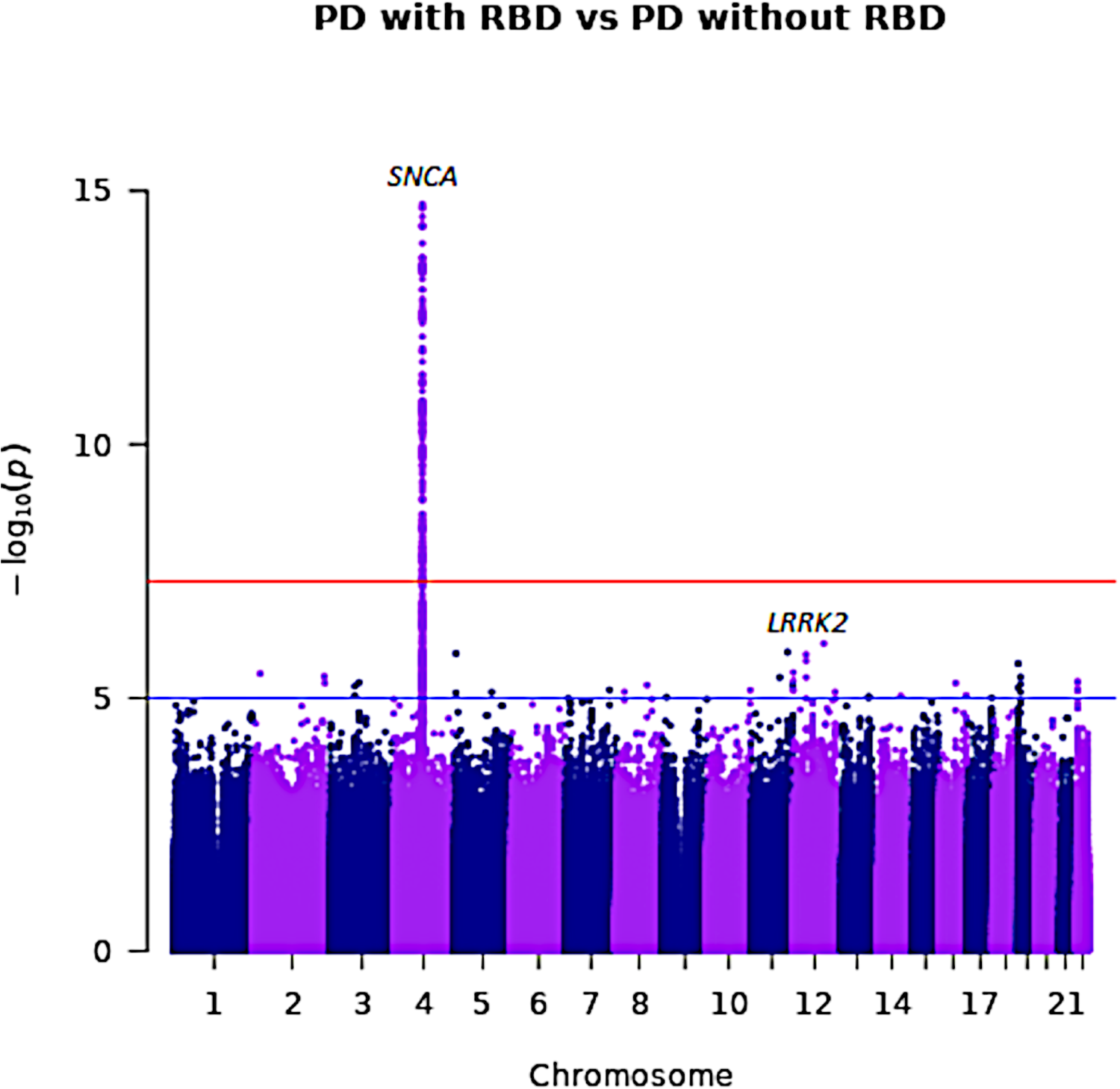
Manhattan plot of PD+RBD vs. PD-RBD. Manhattan plot showing the results of the GWAS meta-analysis, comparing PD+RBD and PD-RBD, highlighting the *SNCA* and *LRRK2* loci. The Y axis represents the negative logarithm of p-value, the X axis represents the chromosomal position of the variants and each dot on the figure represents a SNP. The red line represents the genome-wide Bonferroni-corrected statistical significance threshold (5×10^-8^), whereas the blue line is the false-discovery rate-corrected significance threshold (5×10^-5^). Chr: chromosome; PD: Parkinson’s disease; RBD: REM sleep behavior disorder.

**Table 2.** Association of variants from previous PD GWAS with PD+RBD in the current study.

|  |  | PD with and without RBD |  | PD<sup>15, 37</sup> |  |
| --- | --- | --- | --- | --- | --- |
| Variant | Nearest gene | OR (95% CI) | P-value | OR (95% CI) | P-value |
| 4:90636630 | *SNCA* | 0.85 (0.81-0.89) | 2.46E-10* | 1.17 (1.14-1.2) | 1.13E-36 |
| 12:40734202 | *LRRK2* | 0.41 (0.28-0.61) | 1.04E-05* | 11.35 (9.44-13.63) | 3.61E-148 |
| 4:90626111 | *SNCA* | 0.89 (0.84-0.95) | 0.0001365* | 1.32 (1.29-1.35) | 3.89E-154 |
| 16:30977799 | *SETD1A* | 0.92 (0.88-0.97) | 0.001952 | 1.09 (1.07-1.12) | 5.12E-20 |
| 19:2341047 | *SPPL2B* | 1.09 (1.03-1.15) | 0.002506 | 0.93 (0.91-0.95) | 4.18E-10 |
| 17:43798308 | *CRHR1* | 1.19 (1.05-1.36) | 0.008267 | 0.79 (0.75-0.84) | 6.71E-16 |
| 3:28705690 | *LINC00693* | 0.95 (0.9-0.99) | 0.02552 | 1.07 (1.05-1.09) | 8.09E-12 |

PD: Parkinson's disease; RBD: REM sleep behavior disorder; OR (95% CI): odds ratio with relative 95% confidence interval.
\*variant statistically significant after Bonferroni correction ( $\alpha$ / number of variants=0.00054).

**Table 3.**
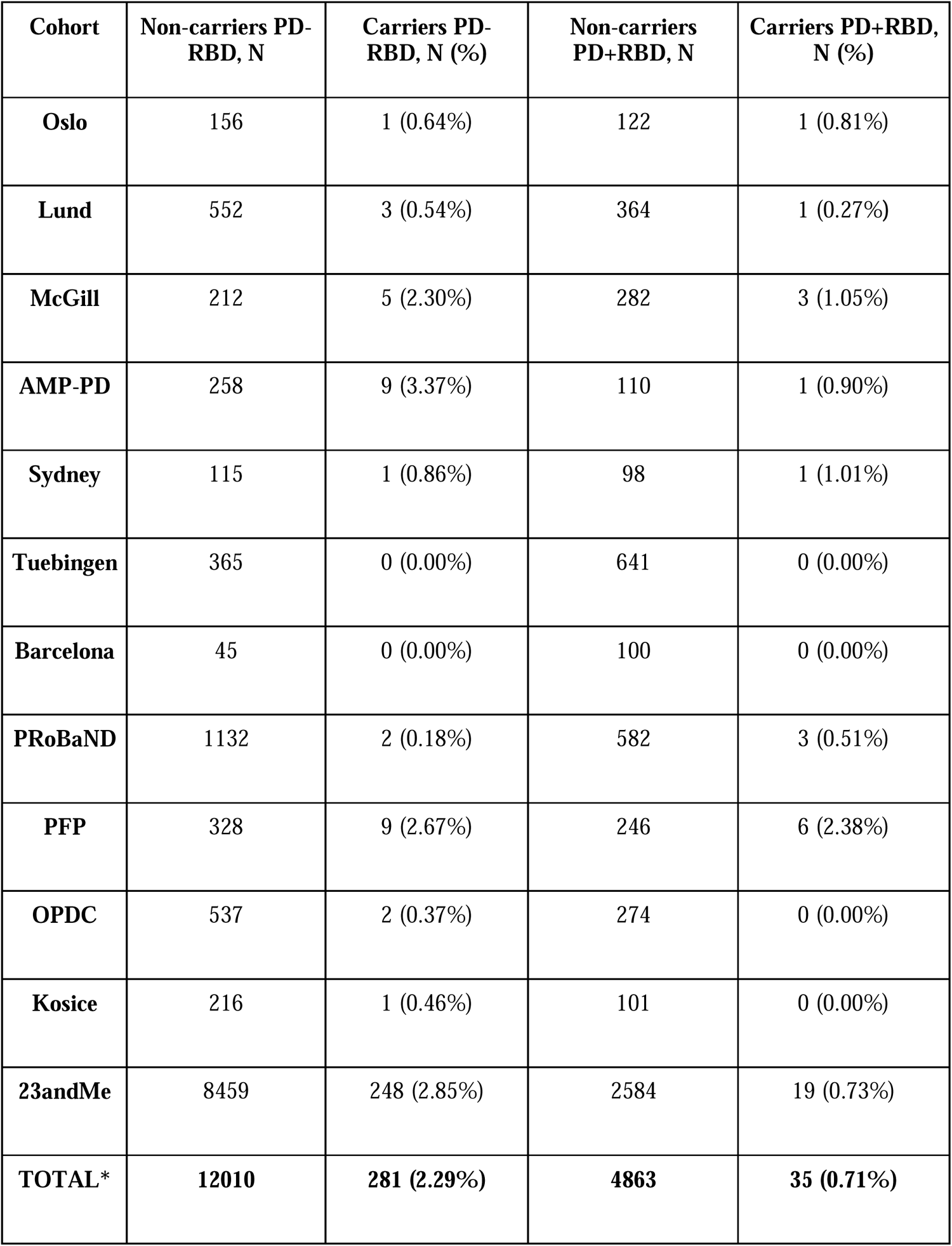
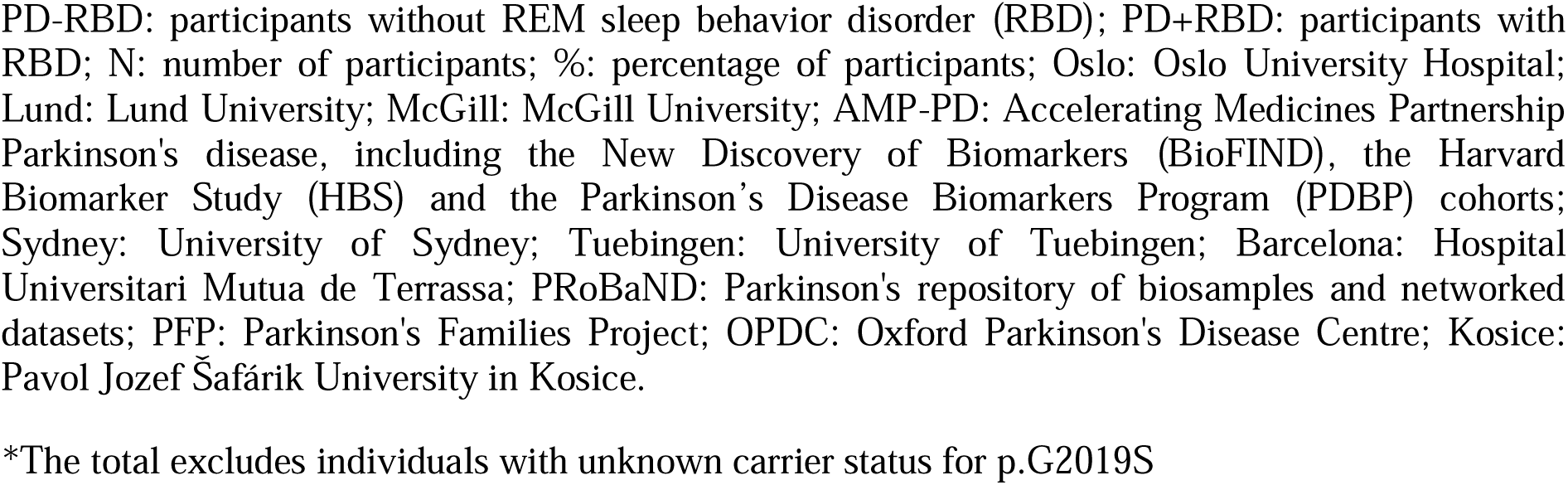
Carriers of the *LRRK2* p.G2019S variant across different cohorts.

| **Cohort** | **Non-carriers PD-RBD, N** | **Carriers PD-RBD, N (%)** | **Non-carriers PD+RBD, N** | **Carriers PD+RBD, N (%)** |
| --- | --- | --- | --- | --- |
| **Oslo** | 156 | 1 (0.64%) | 122 | 1 (0.81%) |
| **Lund** | 552 | 3 (0.54%) | 364 | 1 (0.27%) |
| **McGill** | 212 | 5 (2.30%) | 282 | 3 (1.05%) |
| **AMP-PD** | 258 | 9 (3.37%) | 110 | 1 (0.90%) |
| **Sydney** | 115 | 1 (0.86%) | 98 | 1 (1.01%) |
| **Tuebingen** | 365 | 0 (0.00%) | 641 | 0 (0.00%) |
| **Barcelona** | 45 | 0 (0.00%) | 100 | 0 (0.00%) |
| **PRoBaND** | 1132 | 2 (0.18%) | 582 | 3 (0.51%) |
| **PFP** | 328 | 9 (2.67%) | 246 | 6 (2.38%) |
| **OPDC** | 537 | 2 (0.37%) | 274 | 0 (0.00%) |
| **Kosice** | 216 | 1 (0.46%) | 101 | 0 (0.00%) |
| **23andMe** | 8459 | 248 (2.85%) | 2584 | 19 (0.73%) |
| **TOTAL\*** | **12010** | **281 (2.29%)** | **4863** | **35 (0.71%)** |

PD-RBD: participants without REM sleep behavior disorder (RBD); PD+RBD: participants with RBD; N: number of participants; %: percentage of participants; Oslo: Oslo University Hospital; Lund: Lund University; McGill: McGill University; AMP-PD: Accelerating Medicines Partnership Parkinson's disease, including the New Discovery of Biomarkers (BioFIND), the Harvard Biomarker Study (HBS) and the Parkinson's Disease Biomarkers Program (PDBP) cohorts; Sydney: University of Sydney; Tuebingen: University of Tuebingen; Barcelona: Hospital Universitari Mutua de Terrassa; PRoBaND: Parkinson's repository of biosamples and networked datasets; PFP: Parkinson's Families Project; OPDC: Oxford Parkinson's Disease Centre; Kosice: Pavol Jozef Šafárik University in Kosice.
\*The total excludes individuals with unknown carrier status for p.G2019S

### Genetic correlation and causative associations between PD with RBD and neuropsychiatric disorders

To examine potential genetic correlations between the risk of RBD in PD and multiple neuropsychiatric conditions, we performed LDSC (Fig 2, Supplementary Table 3). We found that the PD+RBD trait was mildly correlated with attention deficit hyperactivity disorder (ADHD, rg=0.30, SE=0.14, *p=*0.04). The most recently published European PD GWAS was genetically correlated with PD-RBD (rg=-0.38, SE=0.15, *p=*0.01). However, these correlations were not statistically significant after Bonferroni correction (α=0.0025).

**Fig 2.**
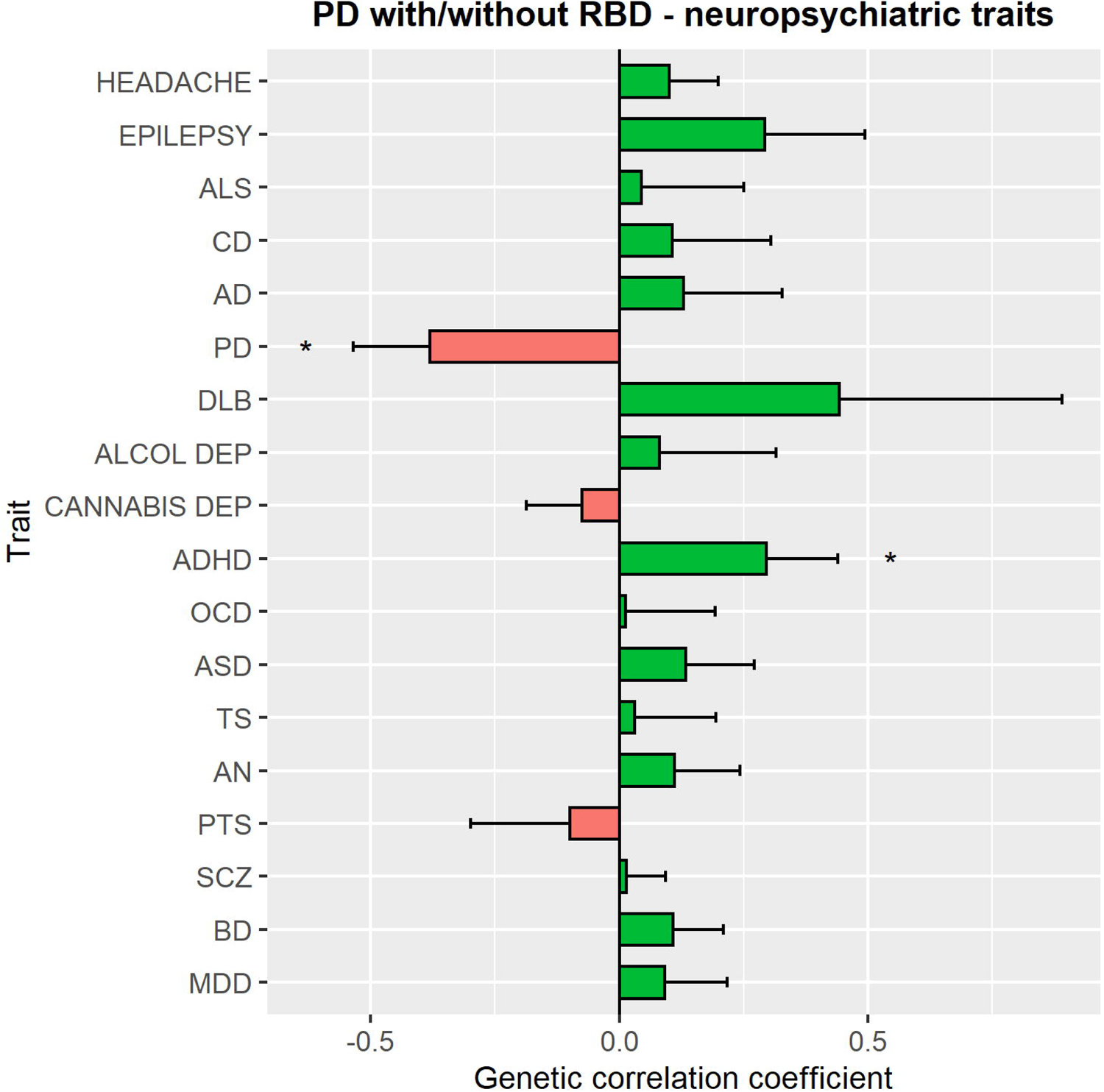
Genetic correlation between PD+RBD and neuropsychiatric traits. The bar plot shows the genetic correlations between PD+RBD and neuropsychiatric traits. The correlation coefficient is illustrated on the X axis. Green bars represent positive correlations whereas red bars negative ones (i.e., a positive correlation of the neuropsychiatric trait with PD-RBD). The asterisks highlight the nominally significant correlations. ALS: amyotrophic lateral sclerosis; CD: cognitive decline; AD: Alzheimer’s disease; PD: Parkinson’s disease; DLB: dementia with Lewy bodies; Alcohol dep: alcohol dependence; cannabis dep: cannabis dependence; ADHD: attention deficit hyperactivity disorder; OCD: obsessive-compulsive disorder; ASD: autism spectrum disorder; TS: Tourette syndrome; AN: anorexia nervosa; PTS: post-traumatic syndrome; scz: schizophrenia; BD: bipolar disorder; MDD: major depressive disorder.

To assess possible causative associations between neuropsychiatric conditions and PD+RBD we performed MR using neuropsychiatric disorders as exposures and PD+RBD as the outcome (Supplementary Figs 2 and 3, Supplementary Tables 4-7). No test showed a statistically significant causative association between neuropsychiatric traits and PD+RBD. However, our power for this analysis was suboptimal (35.7%), therefore there could be associations that we could not detect. We were not able to conduct reverse MR using PD+RBD as the exposure since only one locus passed GWAS significance, preventing us from performing appropriate sensitivity analyses.

## DISCUSSION

In the current GWAS, we found that variants in the *SNCA* and *LRRK2* loci may modify the risk of RBD in PD. Additional loci (*SETD1A*, *SPPL2B*, *CRHR1* and *LINC00693*) require further studies to examine whether they have a role in PD+RBD. The top variant in the *SNCA* locus, rs10005233, was previously reported to be associated with iRBD in a candidate gene study.^14^ Another study, using the Oslo and Parkinson’s Progression Marker Initiative cohorts, found another variant in the *SNCA* locus associated with PD+RBD (rs3756063), which is in strong linkage disequilibrium (LD) with rs10005233 (D’=0.97, r^2^= 0.91).^44^ Furthermore, rs10005233 is in LD with other 5’ region *SNCA* variants associated with synucleinopathies, including rs7681440 (D’=0.99, r^2^=0.94), associated with DLB,^45^ rs763443 (D’=0.89, r^2^=0.78), a secondary PD GWAS signal,^14, 15, 46^ as well as rs2583988 (D’=0.99, r^2^=0.40), a variant located in the *SNCA-AS1* region (discussed below) and associated with Lewy body variant of Alzheimer’s disease (ADLBV).^47^ It is still unclear whether it is a specific variant in the *SNCA* locus or the presence of a specific *SNCA* haplotype that drives these associations with cognitive phenotypes across synucleinopathies.^14^ The rs10005233 variant is also in LD (D’=0.97, r^2^=0.91) with the top signal of a recently published RBD GWAS, rs3756059,^20^ which was associated with reduced expression of *SNCA-AS1,* an antisense RNA molecule that could potentially reduce the translation of alpha-synuclein when it is overexpressed or increase the translation of alpha-synuclein when it is down-regulated. Notably, this reduced expression of *SNCA-AS1* is mainly in cortical areas,^20^ thus potentially increasing alpha-synuclein levels and exposing the cerebral cortex to a greater risk of neurodegeneration in carriers of this RBD-associated variant. The latter hypothesis should be tested in relevant animal models. Altogether, these data suggest that, depending on possible region-specific effects, different *SNCA* variants might play different roles in synucleinopathies.

Similar to PD+RBD, other variants could be involved in the development of PD-RBD. We found that three of the 92 PD GWAS signals associated with increased PD risk in Europeans and Asians,^15, 43, 48^ the LRRK2 variant p.G2019S and the *SNCA* variants rs5019538 and rs356182, were all associated with PD-RBD, compared with PD+RBD. The association between p.G2019S and PD-RBD is in line with a previously reported reduced frequency of RBD in PD patients carrying this variant,^49^ and with lack of p.G2019S carriers in about 1,000 iRBD patients in another study.^19^ In addition to a reduced occurrence of RBD, carriers of the p.G2019S LRRK2 variant also present an overall more benign phenotype, including less frequent and milder cognitive decline.^50, 51^ These findings, in addition to the nominal correlation between PD+RBD and the most recent PD GWAS in Europeans^15^ suggest that such GWAS might explain the genetic background of PD-RBD more than it does for PD+RBD.

These findings further support a pathophysiological relationship between the manifestation of RBD in PD and cognitive decline, which is in line with the comorbidity of these two clinical entities. It was hypothesized that this clinical and pathophysiological correlation could reflect the two alternative directions of alpha-synuclein spreading, body-first or brain-first.^52^ In body-first PD, alpha-synuclein pathology may start in the enteric nervous system, whereas in brain-first PD it may arise in the amygdala, entorhinal cortex and substantia nigra. These different neuropathological patterns correspond to two different subgroups of clinical progression. In body-first PD, RBD may manifest before the motor PD symptoms, and cognitive decline occurs faster, whereas in brain-first PD, RBD may occur after the onset of motor PD symptoms, if at all, and cognitive impairment develop more slowly.^52–55^ We can therefore speculate that the *SNCA* rs10005233 variant associated with PD+RBD might also be associated with the body-first subtype of PD, whereas the LRRK2 p.G2019S variant might be associated with the brain-first PD subtype, with less frequent RBD and milder cognitive decline. Since we cannot determine in our data which patients had RBD prior to PD diagnosis and which had it after PD diagnosis, this hypothesis should be studied in future genetic analyses of brain-first vs. body-first PD.

Similar to previous reports in iRBD and PD+RBD,^14, 15, 44^ in this study we did not observe any involvement of *APOE* variants in PD+RBD, suggesting that this gene does not affect RBD risk in PD patients. The rs117615688 variant (chromosomal position 17:43798308) in the *CRHR1* gene, located in the *MAPT* locus, was nominally associated with RBD (OR=1.19, 95% CI=1.05-1.36, *p*=0.008) with an opposite direction of effect to that seen in PD (OR=0.79, 95% CI=0.75-0.84, *p*=6.71E-16) (Table 2).

There are several limitations in this study. All participants were Europeans, therefore our results might not fully apply to other ancestries. In addition, although we included a large number of patients with PD, insufficient power in our analysis might explain the lack of causative associations between PD+RBD and neuropsychiatric traits as well as of genome-wide significance of the LRRK2 p.G2019S and *GBA1* variants. It is possible that *GBA1* variants are strongly implicated also in the PD subtype without RBD, thus counterbalancing their previously reported contribution to RBD risk.^12, 20^ Another limitation is represented by the inclusion of patients who developed RBD both before and after PD, as they may represent body-first vs. brain-first subtypes of PD as discussed above. Future research with larger sample sizes could investigate possible genetic and biological differences between them and specifically differentiate brain-first and body-first PD in that sense.

In conclusion, in this study we demonstrated that the risk of PD+RBD may be modified by variants in the *SNCA* and *LRRK2* loci, and potentially other loci. These genetic associations may explain why cognitive decline is more frequently observed in PD+RBD compared to PD-RBD, with possible implications for therapeutic management of PD patients. Future research will need to further explore the relationship between genetics, biology and clinical comorbidities to define PD subtypes and implement a precision medicine guided by early markers.

### Relevant conflicts of interest

ZGO has received consulting fees from Lysosomal Therapeutics Inc., Idorsia, Prevail Therapeutics, Denali, Ono Therapeutics, Neuron23, Handl Therapeutics, UBC, Bial Biotech Inc., Bial, Deerfield, Guidepoint, Lighthouse and Inception Sciences (now Ventus). None of these companies were involved in any parts of preparing, drafting and publishing this study. KH, PF, LNK, and the 23andMe Research Team are employed by and hold stock or stock options in 23andMe. HM is employed by UCL. In the last 12 months he reports paid consultancy from Roche, Aprinoia and Amylyx; lecture fees/honoraria - BMJ, Kyowa Kirin, Movement Disorders Society. Research Grants from Parkinson’s UK, Cure Parkinson’s Trust, PSP Association, Medical Research Council, Michael J Fox Foundation. Dr Morris is a co-applicant on a patent application related to C9ORF72 - Method for diagnosing a neurodegenerative disease (PCT/GB2012/052140)

Remaining authors declare no competing interests.

## Data availability

The PD with and without RBD GWAS summary statistics without the 23andMe cohort is publicly available on GWAS catalog (https://www.ebi.ac.uk/gwas/). To access the full summary statistics with 23andMe an application to 23andMe is required. The full GWAS summary statistics for the 23andMe dataset will be made available through 23andMe to qualified researchers under an agreement with 23andMe that protects the privacy of the 23andMe participants. Please visit https://research.23andme.com/collaborate/#dataset-access/ for more information and to apply to access the data. The top 10,000 SNPs GWAS summary statistics including 23andMe is available on https://github.com/gan-orlab. The GWAS summary statistics for the neuropsychiatric traits used in the study are available on the GWAS catalog (https://www.ebi.ac.uk/gwas/) and Psychiatric Genomics Consortium (https://pgc.unc.edu/).

## Code availability

The codes used for the analyses are available on https://github.com/daskrohn/RBD_GWAS and https://github.com/gan-orlab.

## Supporting information

Q-Q plot of the PD with and without RBD GWAS

Leave-one-out analyses results for Mendelian randomization

Single SNP analyses results for Mendelian randomization

PD GWAS hits in the PD with and without RBD GWAS compared with the previous PD GWAS in Europeans/Asians

Carriers of the main GBA1 pathogenic variants across different cohorts

Genetic correlation between PD with RBD and neuropsychiatric traits

Mendelian Randomization results

## Data Availability

All data produced in the present study are available upon reasonable request to the authors. The codes used for the analyses are available on https://github.com/daskrohn/RBD_GWAS and https://github.com/gan-orlab.

https://mcgill-my.sharepoint.com/personal/yuri_sosero_mail_mcgill_ca/_layouts/15/onedrive.aspx?id=%2Fpersonal%2Fyuri%5Fsosero%5Fmail%5Fmcgill%5Fca%2FDocuments%2FSupplementary%20Table%201%2Egz&parent=%2Fpersonal%2Fyuri%5Fsosero%5Fmail%5Fmcgill%5Fca%2FDocuments&ga=1

## Acknowledgement

We wholeheartedly thank the participants in this study. We would like to thank the research participants and all members of IPDGC for making this work possible. The AMP-PD cohort data used in this study included the Fox Investigation for New Discovery of Biomarkers (BioFIND), the Harvard Biomarker Study (HBS) and the Parkinson’s Disease Biomarkers Program (PDBP) cohorts. For up-to-date information on the study, visit https://www.amp-pd.org. AMP PD – a public-private partnership – is managed by the FNIH and funded by Celgene, GSK, the Michael J. Fox Foundation for Parkinson’s Research, the National Institute of Neurological Disorders and Stroke, Pfizer, Sanofi, and Verily. BioFIND is sponsored by The Michael J. Fox Foundation for Parkinson’s Research (MJFF) with support from the National Institute for Neurological Disorders and Stroke (NINDS). The BioFIND Investigators have not participated in reviewing the data analysis or content of the manuscript. For up-to-date information on the study, visit https://www.michaeljfox.org/news/biofind. The Harvard Biomarker Study (HBS) is a collaboration of HBS investigators [full list of HBS investigators found at https://www.bwhparkinsoncenter.org/biobank/ and funded through philanthropy and NIH and Non-NIH funding sources. The HBS Investigators have not participated in reviewing the data analysis or content of the manuscript. The Parkinson’s Disease Biomarker Program (PDBP) consortium is supported by the National Institute of Neurological Disorders and Stroke (NINDS) at the National Institutes of Health. A full list of PDBP investigators can be found at https://pdbp.ninds.nih.gov/policy. The PDBP investigators have not participated in reviewing the data analysis or content of the manuscript.

The PRoBaND cohort is primarily funded and supported by Parkinson’s UK (https://www.parkinsons.org.uk/) and supported by the National Institute for Health Research (NIHR) Dementias and Neurodegenerative Diseases Research Network (DeNDRoN) and by NHS Greater Glasgow and Clyde. PRoBaND has multicentre research ethics approval from the West of Scotland Research Ethics Committee: IRAS 70980, MREC 11/AL/0163. The Oxford Parkinson’s Disease Centre (OPDC) Discovery cohort is also primarily funded by Parkinson’s UK and additionally supported by the NIHR-DeNDRoN and by the NIHR Oxford Biomedical Research Centre, based at the Oxford University Hospitals NHS Trust, and the University of Oxford. OPDC has multicentre research ethics approval from the South Central Oxford A Research Ethics Committee: 16/SC/0108. The Parkinson’s Family Project (PFP) cohort has received funding from Parkinson’s UK and the Janet Owens Bequest Fund. PFP has multicentre research ethics approval from the London – Camden and Kings Cross Research Ethics Committee: 5/LO/0097. RR is funded by Aligning Science Across Parkinson’s (grant number ASAP-000478) through the Michael J. Fox Foundation for Parkinson’s Research (MJFF). This work was financially supported by the Michael J. Fox Foundation, Parkinson’s Society Canada, the Canadian Consortium on Neurodegeneration in Aging (CCNA), and the Canada First Research Excellence Fund (CFREF), awarded to McGill University for the Healthy Brains for Healthy Lives (HBHL) program. ZGO is supported by the Fonds de recherche du Québec - Santé (FRQS) Chercheurs-boursiers award, and is a William Dawson Scholar.

We would also like to thank the research participants and employees of 23andMe for making this work possible.

The following members of the 23andMe Research Team contributed to this study:

Stella Aslibekyan, Adam Auton, Elizabeth Babalola, Robert K. Bell, Jessica Bielenberg, Jonathan Bowes, Katarzyna Bryc, Ninad S. Chaudhary, Daniella Coker, Sayantan Das, Emily DelloRusso, Sarah L. Elson, Nicholas Eriksson, Teresa Filshtein, Pierre Fontanillas, Will Freyman, Zach Fuller, Chris German, Julie M. Granka, Karl Heilbron, Alejandro Hernandez, Barry Hicks, David A. Hinds, Ethan M. Jewett, Yunxuan Jiang, Katelyn Kukar, Alan Kwong, Yanyu Liang, Keng-Han Lin, Bianca A. Llamas, Matthew H. McIntyre, Steven J. Micheletti, Meghan E. Moreno, Priyanka Nandakumar, Dominique T. Nguyen, Jared O’Connell, Aaron A. Petrakovitz, G. David Poznik, Alexandra Reynoso, Shubham Saini, Morgan Schumacher, Leah Selcer, Anjali J. Shastri, Janie F. Shelton, Jingchunzi Shi, Suyash Shringarpure, Qiaojuan Jane Su, Susana A. Tat, Vinh Tran, Joyce Y. Tung, Xin Wang, Wei Wang, Catherine H. Weldon, Peter Wilton, Corinna D. Wong.

K.H., P.F., L.K., and 23andMe Research Team are employed by and hold stock or stock options in 23andMe, Inc.

## Author contributions

YLS, ZGO, LK: conception and design of the study

YLS, LK, KH, PF, LNK, EY, UR, JAR, KF, FA, KB, MS, HM, LW, RR, LP, MT, TG, KB, HL, MTMH, DGG, SJGL, JBK, PP, IA, MS, AL, MO, MR, ZGO: data acquisition/analysis

YLS, ZGO: manuscript drafting

---

**Supplementary Fig 1 Q-Q plot of the PD with and without RBD GWAS**

The Q-Q plot illustrates the negative log-adjusted p-values from the GWAS sorted into ascending order against the expected quantiles if the null hypothesis is true for all tests.

**Supplementary Fig 2 Leave-one-out analysis in the Mendelian Randomization**

The plot shows the Mendelian Randomization coefficient (black dot) and 95% confidence interval (bars) when each variant is excluded from the analysis. The excluded variant is indicated on the Y axis. The red dot and line represent the results if all the variants are included.

SNP: single nucleotide polymorphism. MR: Mendelian Randomization; PD: Parkinson’s disease; RBD: REM sleep behavior disorder.

**Supplementary Fig 3 Single variant analysis in the Mendelian Randomization**

The plot shows the Mendelian Randomization coefficient (black dot) and 95% confidence interval (bars) for each individual variant included in the analysis. The variant analyzed is indicated on the Y axis. The red dot and line represent the results if all the variants are included.

SNP: single nucleotide polymorphism. MR: Mendelian Randomization; PD: Parkinson’s disease; RBD: REM sleep behavior disorder.

## Notes

### Author Declarations

Ethics committee/IRB of McGill University gave ethical approval for this work.

## REFERENCES

1. St Louis EK, Boeve BF. REM Sleep Behavior Disorder: Diagnosis, Clinical Implications, and Future Directions. Mayo Clin Proc. 2017 Nov;92(11):1723–36.

2. Högl B, Stefani A, Videnovic A. Idiopathic REM sleep behaviour disorder and neurodegeneration - an update. Nat Rev Neurol. 2018 Jan;14(1):40–55.

3. Postuma RB, Iranzo A, Hu M, et al. Risk and predictors of dementia and parkinsonism in idiopathic REM sleep behaviour disorder: a multicentre study. Brain. 2019 Mar 1;142(3):744–59.

4. Postuma RB, Gagnon JF, Vendette M, Fantini ML, Massicotte-Marquez J, Montplaisir J. Quantifying the risk of neurodegenerative disease in idiopathic REM sleep behavior disorder. Neurology. 2009 Apr 14;72(15):1296–300.

5. Ayers JI, Lee J, Monteiro O, et al. Different α-synuclein prion strains cause dementia with Lewy bodies and multiple system atrophy. Proc Natl Acad Sci U S A. 2022 Feb 8;119(6).

6. Hu MT. REM sleep behavior disorder (RBD). Neurobiol Dis. 2020 Sep;143:104996.

7. Barasa A, Wang J, Dewey RB, Jr. Probable REM Sleep Behavior Disorder Is a Risk Factor for Symptom Progression in Parkinson Disease. Front Neurol. 2021;12:651157.

8. Diaconu Ș, Falup-Pecurariu O, Țînț D, Falup-Pecurariu C. REM sleep behaviour disorder in Parkinson’s disease (Review). Exp Ther Med. 2021 Aug;22(2):812.

9. Bargiotas P, Ntafouli M, Lachenmayer ML, Krack P, Schüpbach WMM, Bassetti CLA. Apathy in Parkinson’s disease with REM sleep behavior disorder. J Neurol Sci. 2019 Apr 15;399:194–8.

10. Liu Y, Lawton MA, Lo C, et al. Longitudinal Changes in Parkinson’s Disease Symptoms with and Without Rapid Eye Movement Sleep Behavior Disorder: The Oxford Discovery Cohort Study. Mov Disord. 2021 Dec;36(12):2821–32.

11. Duarte Folle A, Paul KC, Bronstein JM, Keener AM, Ritz B. Clinical progression in Parkinson’s disease with features of REM sleep behavior disorder: A population-based longitudinal study. Parkinsonism Relat Disord. 2019 May;62:105–11.

12. Krohn L, Ruskey JA, Rudakou U, et al. GBA variants in REM sleep behavior disorder: A multicenter study. Neurology. 2020 Aug 25;95(8):e1008–e16.

13. Krohn L, Öztürk TN, Vanderperre B, et al. Genetic, Structural, and Functional Evidence Link TMEM175 to Synucleinopathies. Ann Neurol. 2020 Jan;87(1):139–53.

14. Krohn L, Wu RYJ, Heilbron K, et al. Fine-Mapping of SNCA in Rapid Eye Movement Sleep Behavior Disorder and Overt Synucleinopathies. Ann Neurol. 2020 Apr;87(4):584–98.

15. Nalls MA, Blauwendraat C, Vallerga CL, et al. Identification of novel risk loci, causal insights, and heritable risk for Parkinson’s disease: a meta-analysis of genome-wide association studies. Lancet Neurol. 2019 Dec;18(12):1091–102.

16. Chia R, Sabir MS, Bandres-Ciga S, et al. Genome sequencing analysis identifies new loci associated with Lewy body dementia and provides insights into its genetic architecture. Nat Genet. 2021 Mar;53(3):294–303.

17. Ouled Amar Bencheikh B, Ruskey JA, Arnulf I, et al. LRRK2 protective haplotype and full sequencing study in REM sleep behavior disorder. Parkinsonism Relat Disord. 2018 Jul;52:98–101.

18. Gan-Or Z, Montplaisir JY, Ross JP, et al. The dementia-associated APOE ε4 allele is not associated with rapid eye movement sleep behavior disorder. Neurobiol Aging. 2017 Jan;49:218.e13–e15.

19. Mufti K, Rudakou U, Yu E, et al. Comprehensive Analysis of Familial Parkinsonism Genes in Rapid-Eye-Movement Sleep Behavior Disorder. Mov Disord. 2021 Jan;36(1):235–40.

20. Krohn L, Heilbron K, Blauwendraat C, et al. Genome-wide association study of REM sleep behavior disorder identifies polygenic risk and brain expression effects. Nat Commun. 2022 Dec 5;13(1):7496.

21. Postuma RB, Arnulf I, Hogl B, et al. A single-question screen for rapid eye movement sleep behavior disorder: a multicenter validation study. Mov Disord. 2012 Jun;27(7):913–6.

22. Nomura T, Inoue Y, Kagimura T, Uemura Y, Nakashima K. Utility of the REM sleep behavior disorder screening questionnaire (RBDSQ) in Parkinson’s disease patients. Sleep Med. 2011 Aug;12(7):711–3.

23. Skorvanek M, Feketeova E, Kurtis MM, Rusz J, Sonka K. Accuracy of Rating Scales and Clinical Measures for Screening of Rapid Eye Movement Sleep Behavior Disorder and for Predicting Conversion to Parkinson’s Disease and Other Synucleinopathies. Front Neurol. 2018;9:376.

24. Hughes AJ, Daniel SE, Kilford L, Lees AJ. Accuracy of clinical diagnosis of idiopathic Parkinson’s disease: a clinico-pathological study of 100 cases. J Neurol Neurosurg Psychiatry. 1992 Mar;55(3):181–4.

25. Postuma RB, Berg D, Stern M, et al. MDS clinical diagnostic criteria for Parkinson’s disease. Mov Disord. 2015 Oct;30(12):1591–601.

26. Durand EY, Do CB, Mountain JL, Macpherson JM. Ancestry Composition: A Novel, Efficient Pipeline for Ancestry Deconvolution. bioRxiv. 2014:010512.

27. Henn BM, Hon L, Macpherson JM, et al. Cryptic distant relatives are common in both isolated and cosmopolitan genetic samples. PLoS One. 2012;7(4):e34267.

28. Loh PR, Danecek P, Palamara PF, et al. Reference-based phasing using the Haplotype Reference Consortium panel. Nat Genet. 2016 Nov;48(11):1443–8.

29. Browning SR, Browning BL. Rapid and accurate haplotype phasing and missing-data inference for whole-genome association studies by use of localized haplotype clustering. Am J Hum Genet. 2007 Nov;81(5):1084–97.

30. Huang J, Howie B, McCarthy S, et al. Improved imputation of low-frequency and rare variants using the UK10K haplotype reference panel. Nat Commun. 2015 Sep 14;6:8111.

31. Auton A, Brooks LD, Durbin RM, et al. A global reference for human genetic variation. Nature. 2015 Oct 1;526(7571):68–74.

32. Walter K, Min JL, Huang J, et al. The UK10K project identifies rare variants in health and disease. Nature. 2015 Oct 1;526(7571):82–90.

33. Das S, Forer L, Schönherr S, et al. Next-generation genotype imputation service and methods. Nat Genet. 2016 Oct;48(10):1284–7.

34. Purcell S, Neale B, Todd-Brown K, et al. PLINK: a tool set for whole-genome association and population-based linkage analyses. Am J Hum Genet. 2007 Sep;81(3):559–75.

35. Willer CJ, Li Y, Abecasis GR. METAL: fast and efficient meta-analysis of genomewide association scans. Bioinformatics. 2010 Sep 1;26(17):2190–1.

36. Yang J, Ferreira T, Morris AP, et al. Conditional and joint multiple-SNP analysis of GWAS summary statistics identifies additional variants influencing complex traits. Nat Genet. 2012 Mar 18;44(4):369–75, s1-3.

37. Zheng J, Erzurumluoglu AM, Elsworth BL, et al. LD Hub: a centralized database and web interface to perform LD score regression that maximizes the potential of summary level GWAS data for SNP heritability and genetic correlation analysis. Bioinformatics. 2017 Jan 15;33(2):272–9.

38. Smith GD, Ebrahim S. ’Mendelian randomization’: can genetic epidemiology contribute to understanding environmental determinants of disease? Int J Epidemiol. 2003 Feb;32(1):1–22.

39. Burgess S, Small DS, Thompson SG. A review of instrumental variable estimators for Mendelian randomization. Stat Methods Med Res. 2017 Oct;26(5):2333–55.

40. Hemani G, Zheng J, Elsworth B, et al. The MR-Base platform supports systematic causal inference across the human phenome. Elife. 2018 May 30;7.

41. Noyce AJ, Bandres-Ciga S, Kim J, et al. The Parkinson’s Disease Mendelian Randomization Research Portal. Mov Disord. 2019 Dec;34(12):1864–72.

42. Burgess S. Sample size and power calculations in Mendelian randomization with a single instrumental variable and a binary outcome. Int J Epidemiol. 2014 Jun;43(3):922–9.

43. Foo JN, Chew EGY, Chung SJ, et al. Identification of Risk Loci for Parkinson Disease in Asians and Comparison of Risk Between Asians and Europeans: A Genome-Wide Association Study. JAMA Neurol. 2020 Jun 1;77(6):746–54.

44. Bjørnarå KA, Pihlstrøm L, Dietrichs E, Toft M. Risk variants of the α-synuclein locus and REM sleep behavior disorder in Parkinson’s disease: a genetic association study. BMC Neurol. 2018 Feb 21;18(1):20.

45. Guerreiro R, Ross OA, Kun-Rodrigues C, et al. Investigating the genetic architecture of dementia with Lewy bodies: a two-stage genome-wide association study. Lancet Neurol. 2018 Jan;17(1):64–74.

46. Chang D, Nalls MA, Hallgrímsdóttir IB, et al. A meta-analysis of genome-wide association studies identifies 17 new Parkinson’s disease risk loci. Nat Genet. 2017 Oct;49(10):1511–6.

47. Linnertz C, Lutz MW, Ervin JF, et al. The genetic contributions of SNCA and LRRK2 genes to Lewy Body pathology in Alzheimer’s disease. Hum Mol Genet. 2014 Sep 15;23(18):4814–21.

48. Nalls MA, Pankratz N, Lill CM, et al. Large-scale meta-analysis of genome-wide association data identifies six new risk loci for Parkinson’s disease. Nat Genet. 2014 Sep;46(9):989–93.

49. Pont-Sunyer C, Iranzo A, Gaig C, et al. Sleep Disorders in Parkinsonian and Nonparkinsonian LRRK2 Mutation Carriers. PLoS One. 2015;10(7):e0132368.

50. Alcalay RN, Mejia-Santana H, Mirelman A, et al. Neuropsychological performance in LRRK2 G2019S carriers with Parkinson’s disease. Parkinsonism Relat Disord. 2015 Feb;21(2):106–10.

51. Healy DG, Falchi M, O’Sullivan SS, et al. Phenotype, genotype, and worldwide genetic penetrance of LRRK2-associated Parkinson’s disease: a case-control study. Lancet Neurol. 2008 Jul;7(7):583–90.

52. Borghammer P, Horsager J, Andersen K, et al. Neuropathological evidence of body-first vs. brain-first Lewy body disease. Neurobiol Dis. 2021 Dec;161:105557.

53. Horsager J, Knudsen K, Sommerauer M. Clinical and imaging evidence of brain-first and body-first Parkinson’s disease. Neurobiol Dis. 2022 Mar;164:105626.

54. Ferri R, Cosentino FI, Pizza F, Aricò D, Plazzi G. The timing between REM sleep behavior disorder and Parkinson’s disease. Sleep Breath. 2014 May;18(2):319–23.

55. Nomura T, Kishi M, Nakashima K. Differences in clinical characteristics when REM sleep behavior disorder precedes or comes after the onset of Parkinson’s disease. J Neurol Sci. 2017 Nov 15;382:58–60.

