## Supplementary figures and images for "Genome-wide association study of REM sleep behavior disorder in Parkinson’s disease"

### Q-Q plot of the PD with and without RBD GWAS

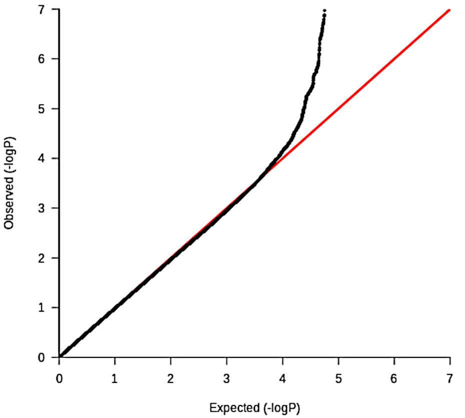

### Single SNP analyses results for Mendelian randomization

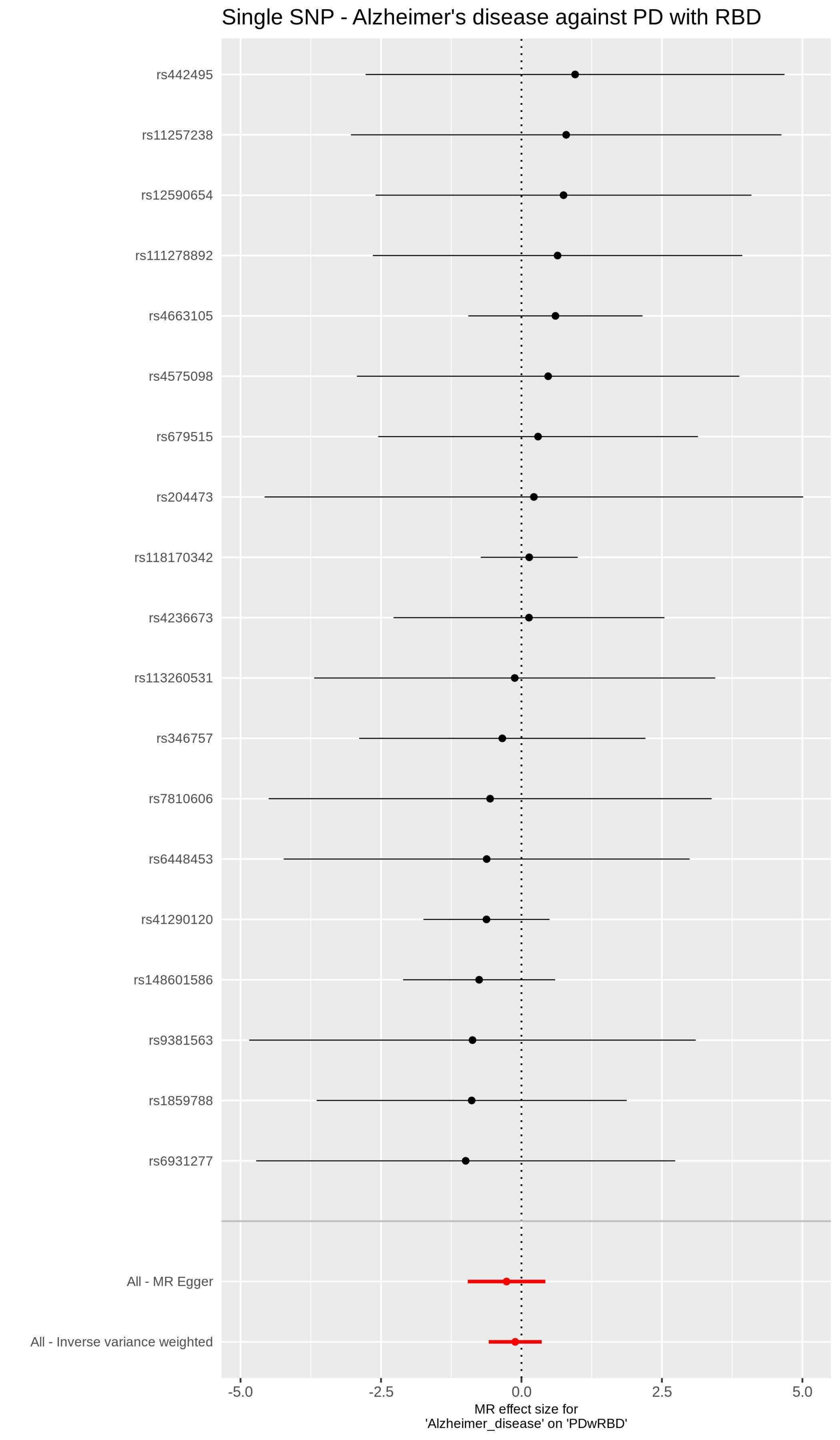

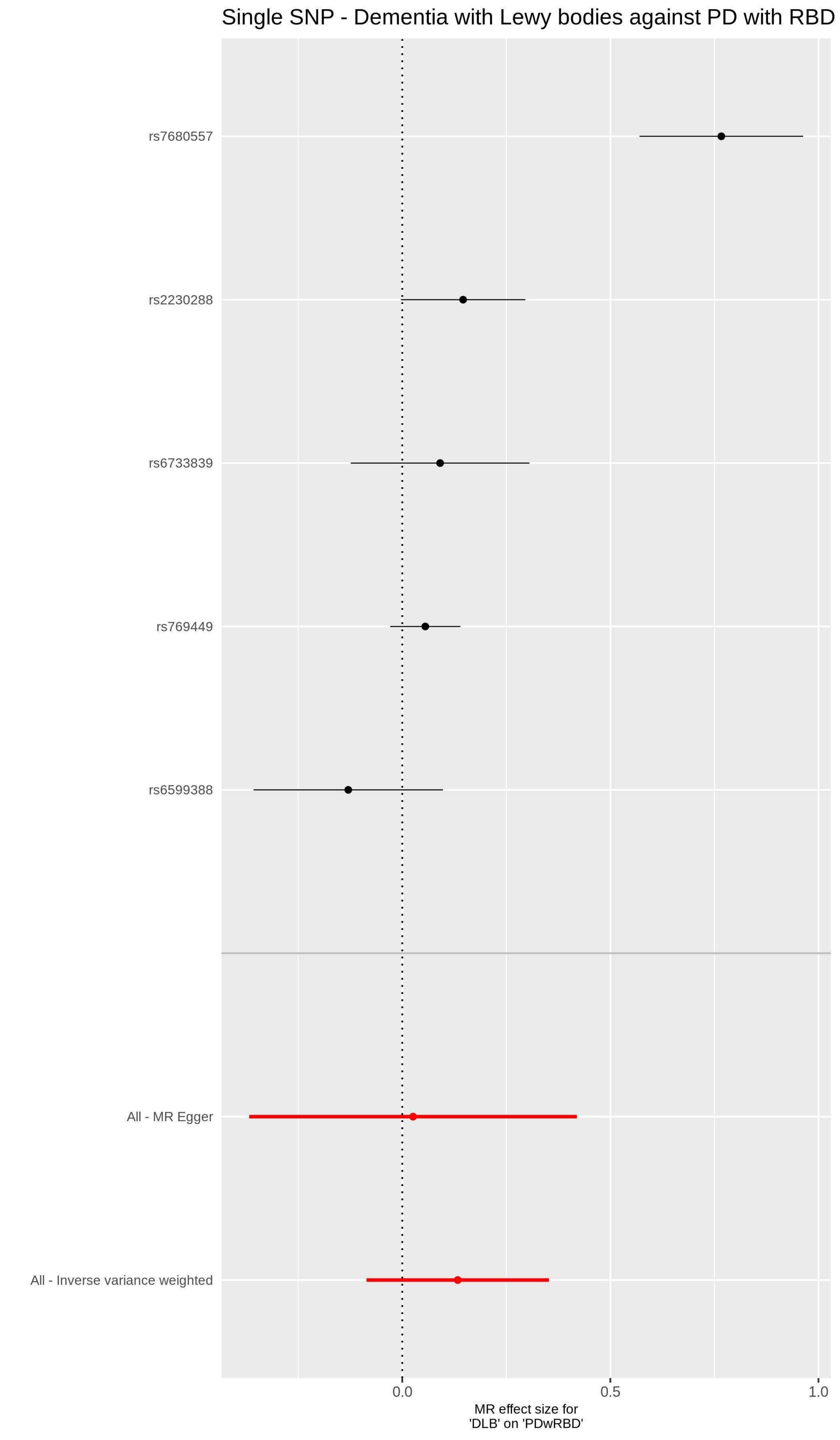

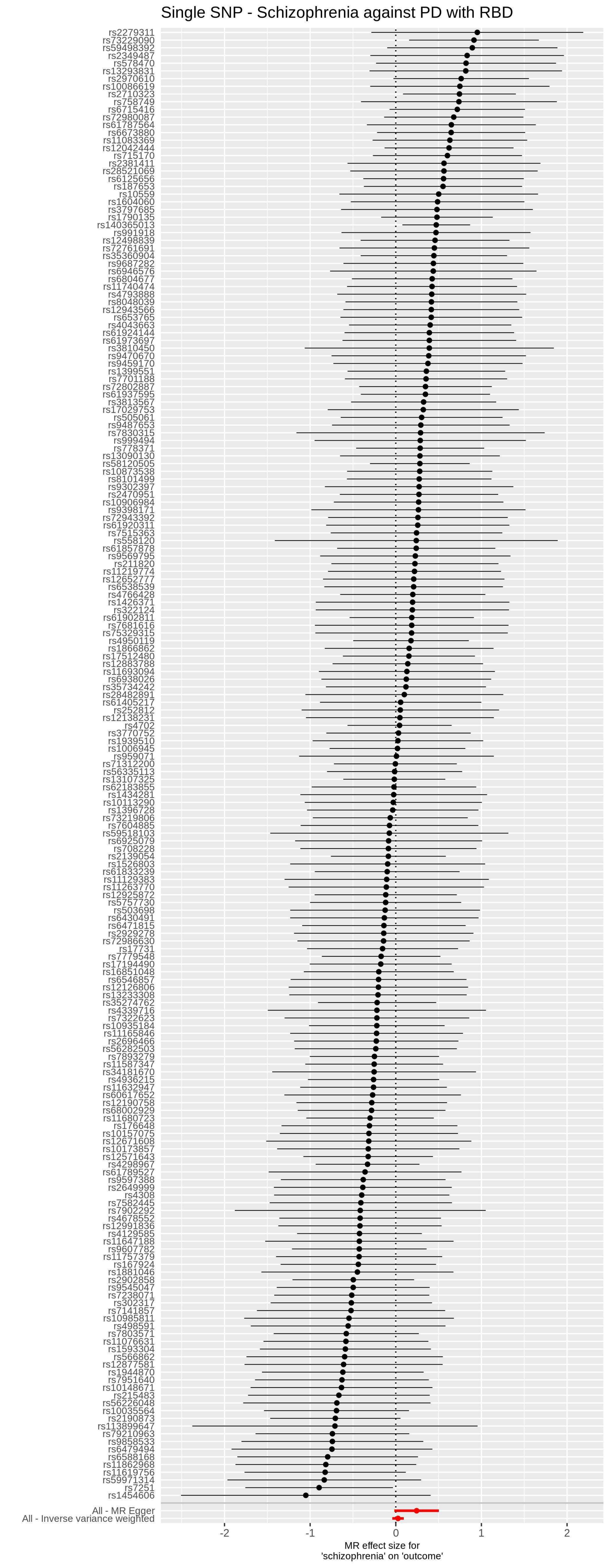

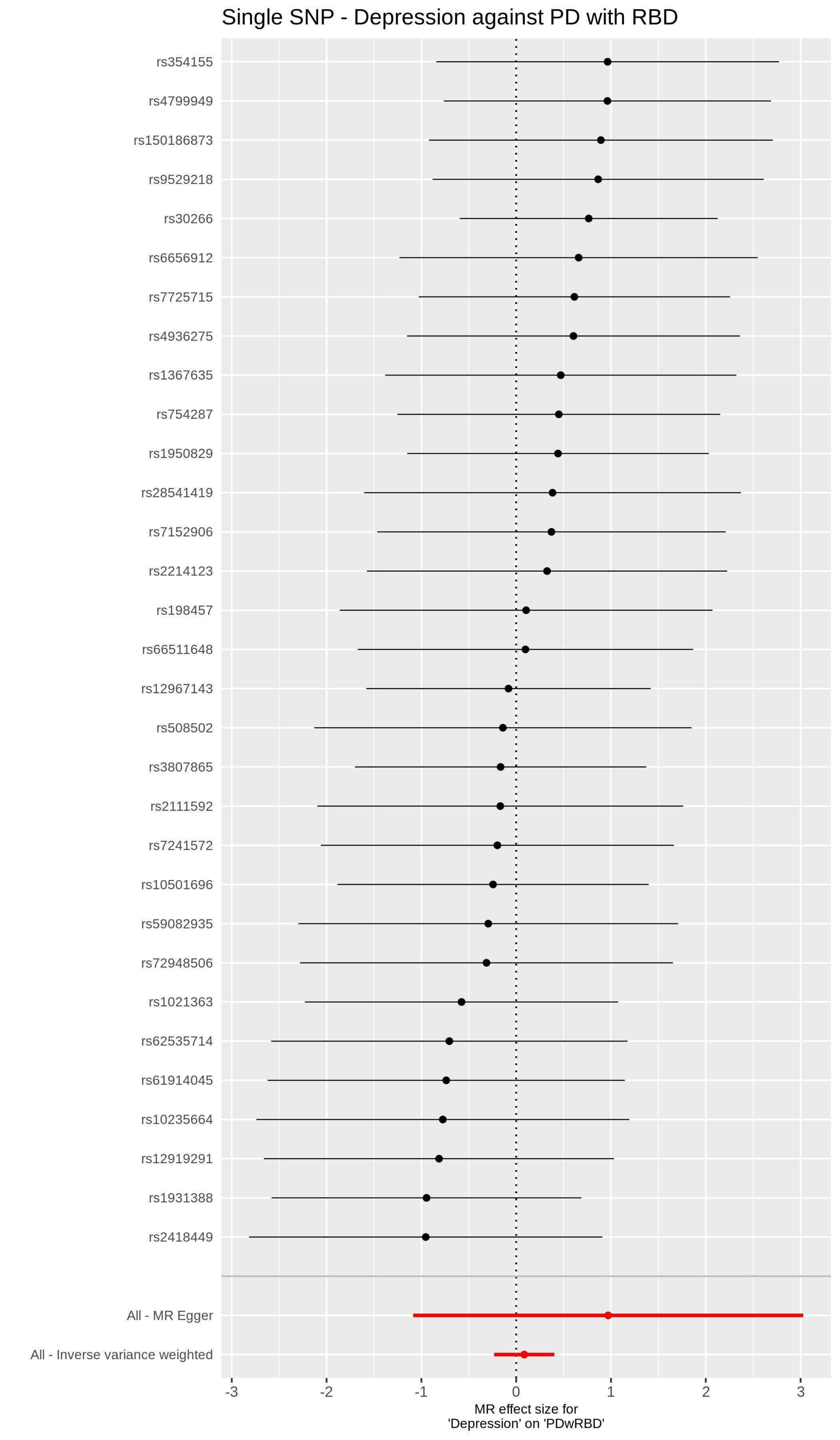

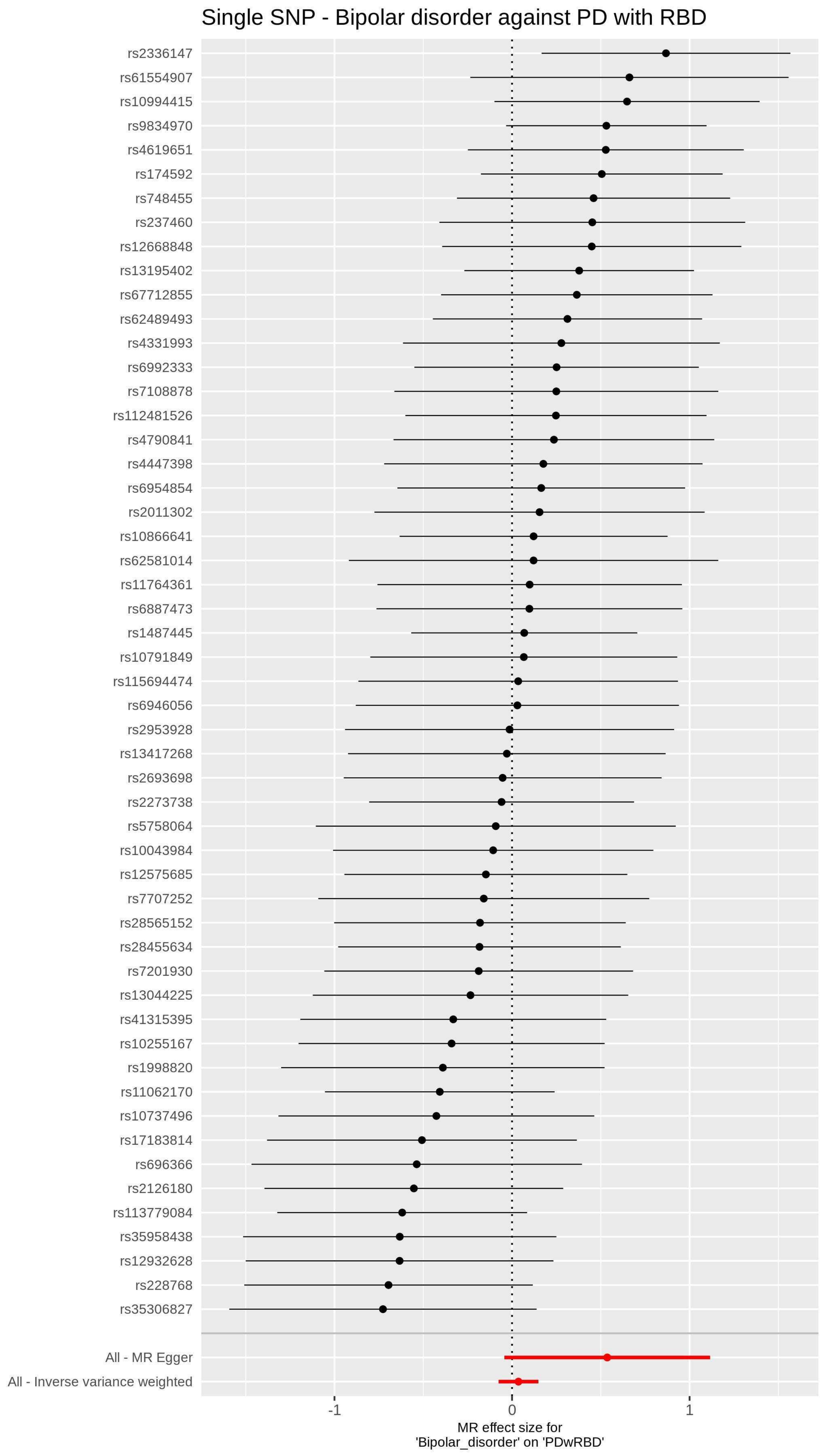
