## Supplementary material for "Genome-wide association study of REM sleep behavior disorder in Parkinson’s disease": Leave-one-out analyses results for Mendelian randomization

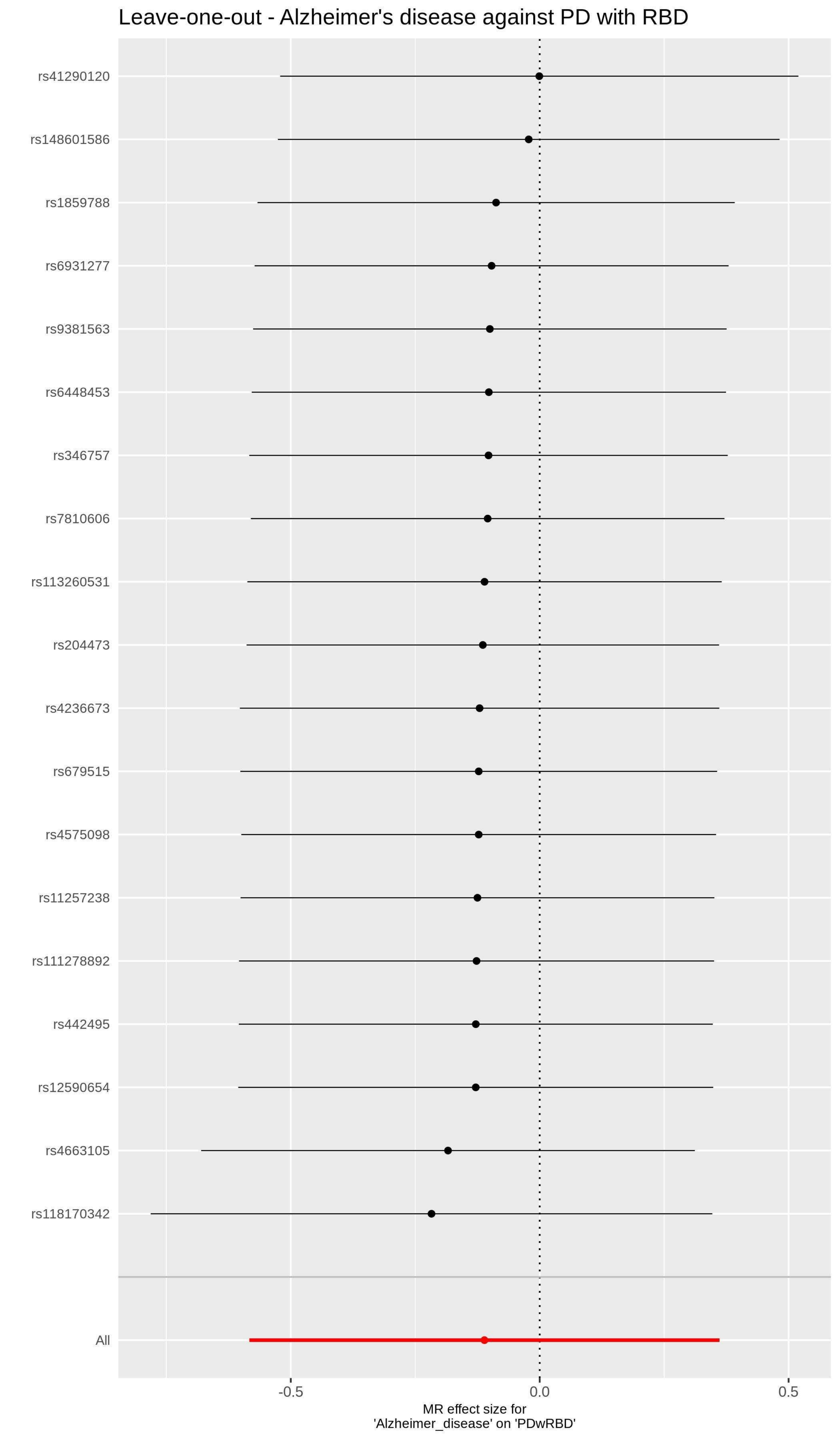
Leave-one-out - Alzheimer's disease against PD with RBD
rs41290120
rs148601586
rs1859788
rs6931277
rs9381563
rs6448453
rs346757
rs7810606
rs113260531
rs204473
rs4236673
rs679515
rs4575098
rs11257238
rs111278892
rs442495
rs12590654
rs4663105
rs118170342
All
-0.5
0.0
0.5
MR effect size for
'Alzheimer_disease' on 'PDwRBD'

Leave-one-out - Dementia with Lewy bodies against PD with RBD

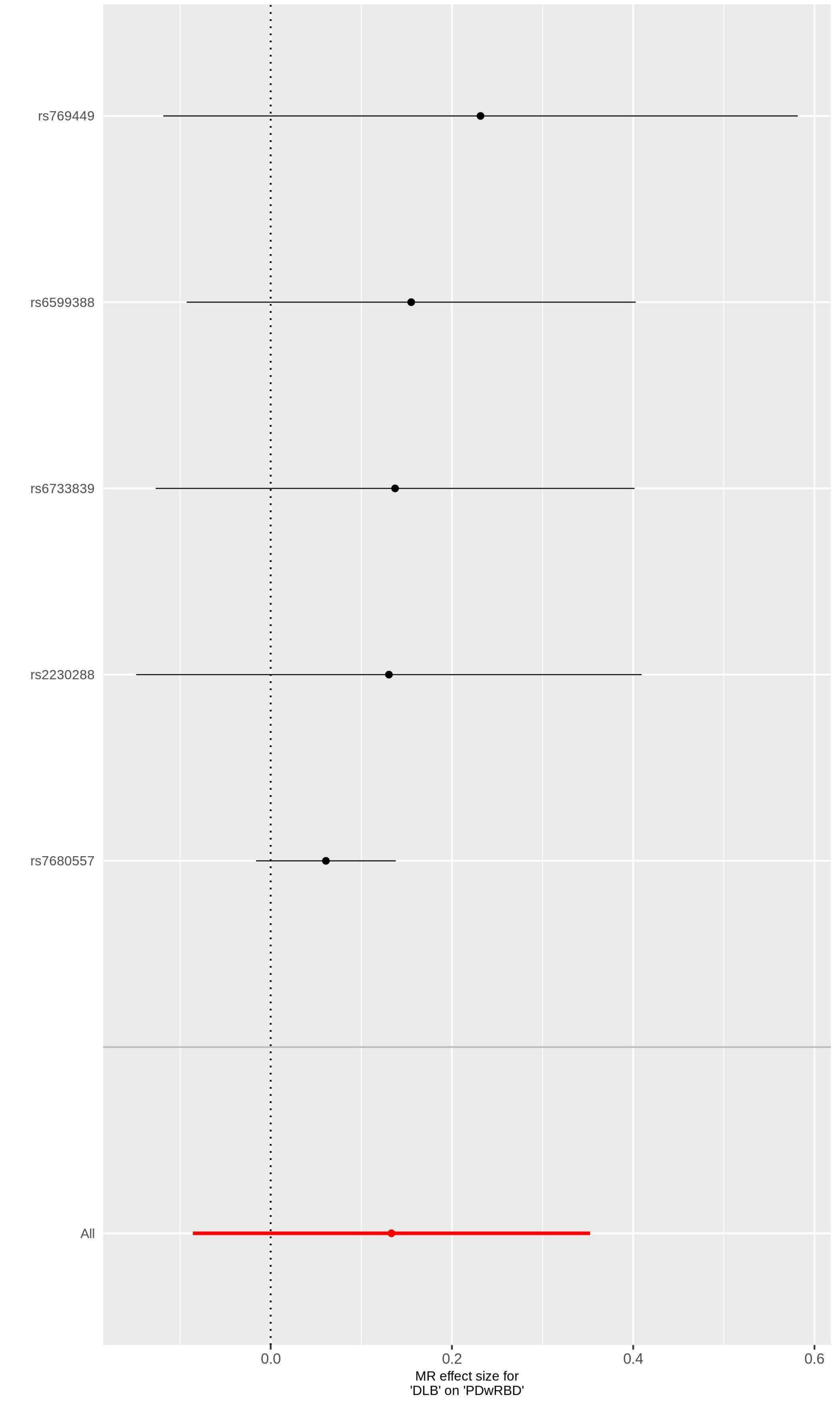

### Leave-one-out - Schizophrenia against PD with RBD

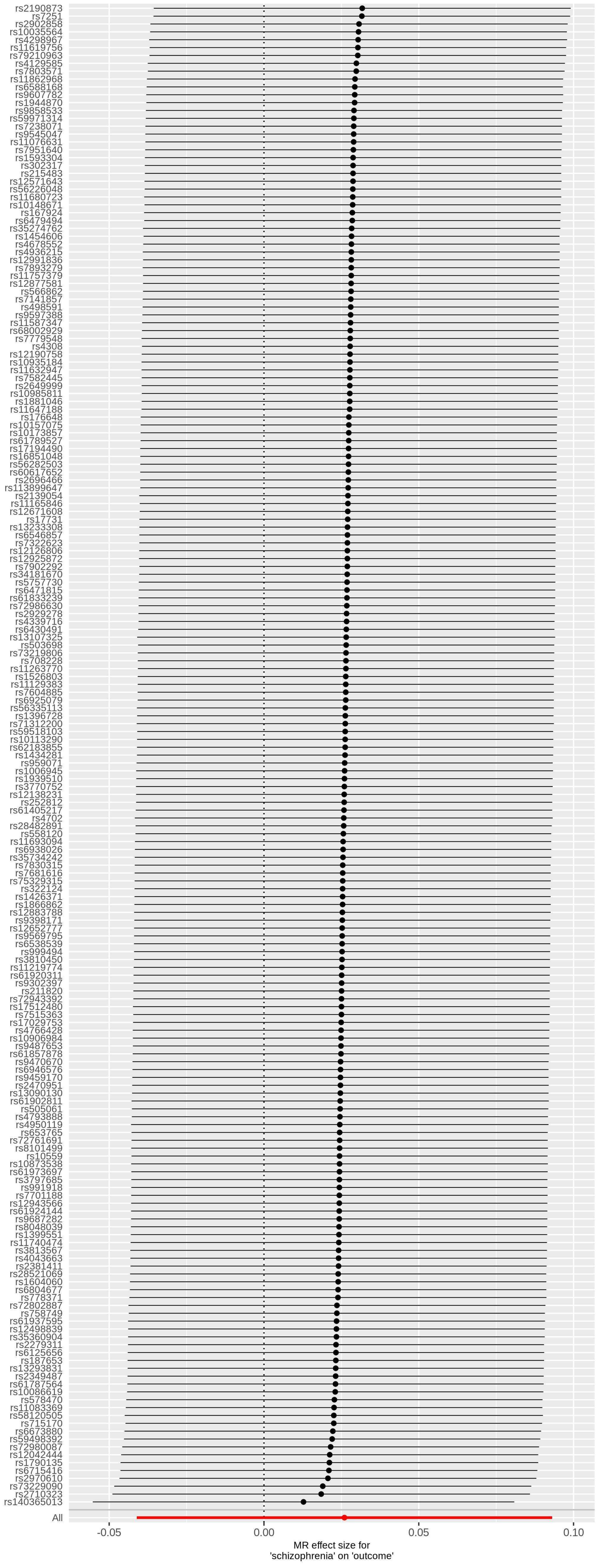

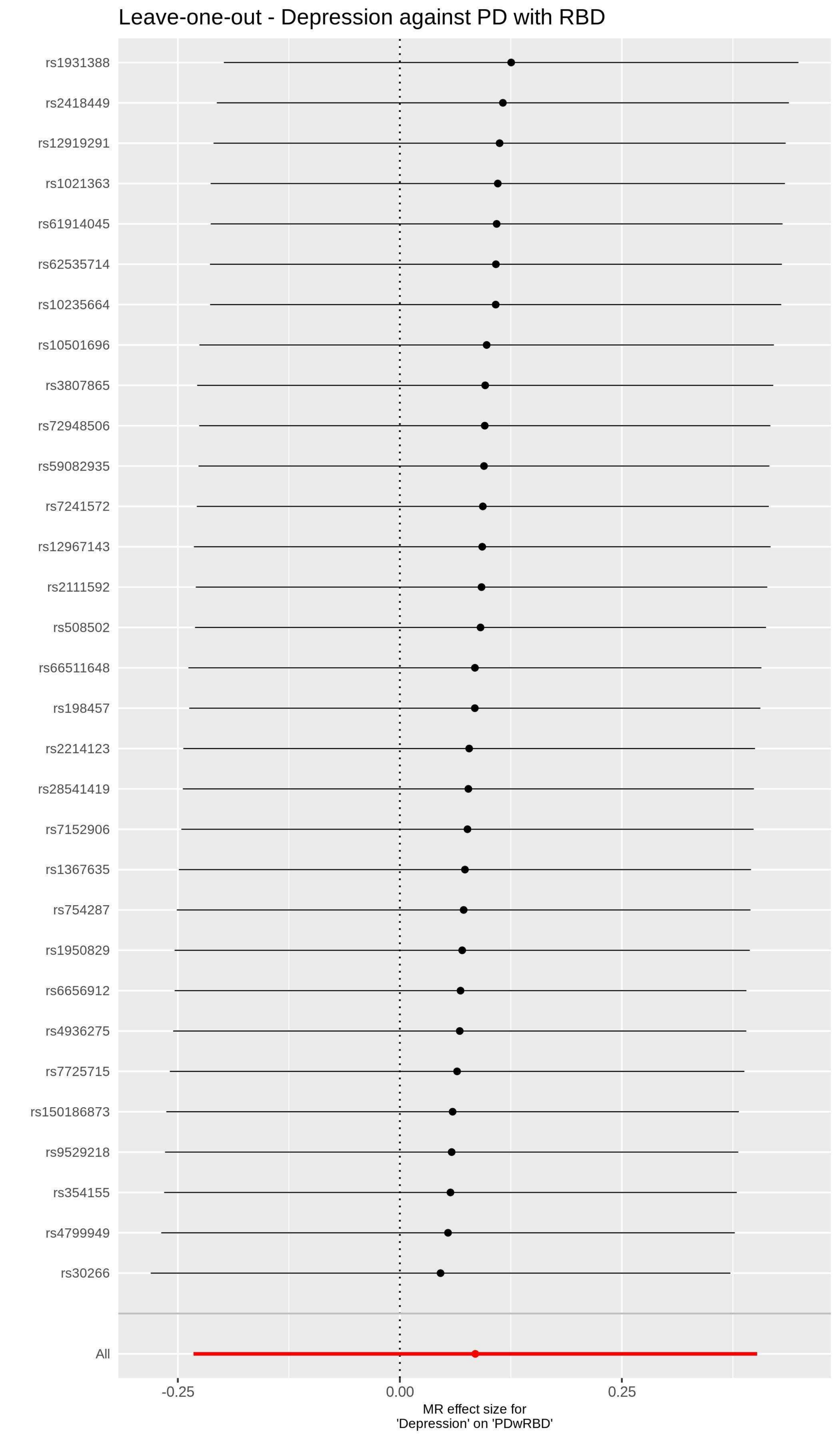

Leave-one-out - Depression against PD with RBD
rs1931388
rs2418449
rs12919291
rs1021363
rs61914045
rs62535714
rs10235664
rs10501696
rs3807865
rs72948506
rs59082935
rs7241572
rs12967143
rs2111592
rs508502
rs66511648
rs198457
rs2214123
rs28541419
rs7152906
rs1367635
rs754287
rs1950829
rs6656912
rs4936275
rs7725715
rs150186873
rs9529218
rs354155
rs4799949
rs30266
All
-0.25
0.00
0.25
MR effect size for
'Depression' on 'PDwRBD'

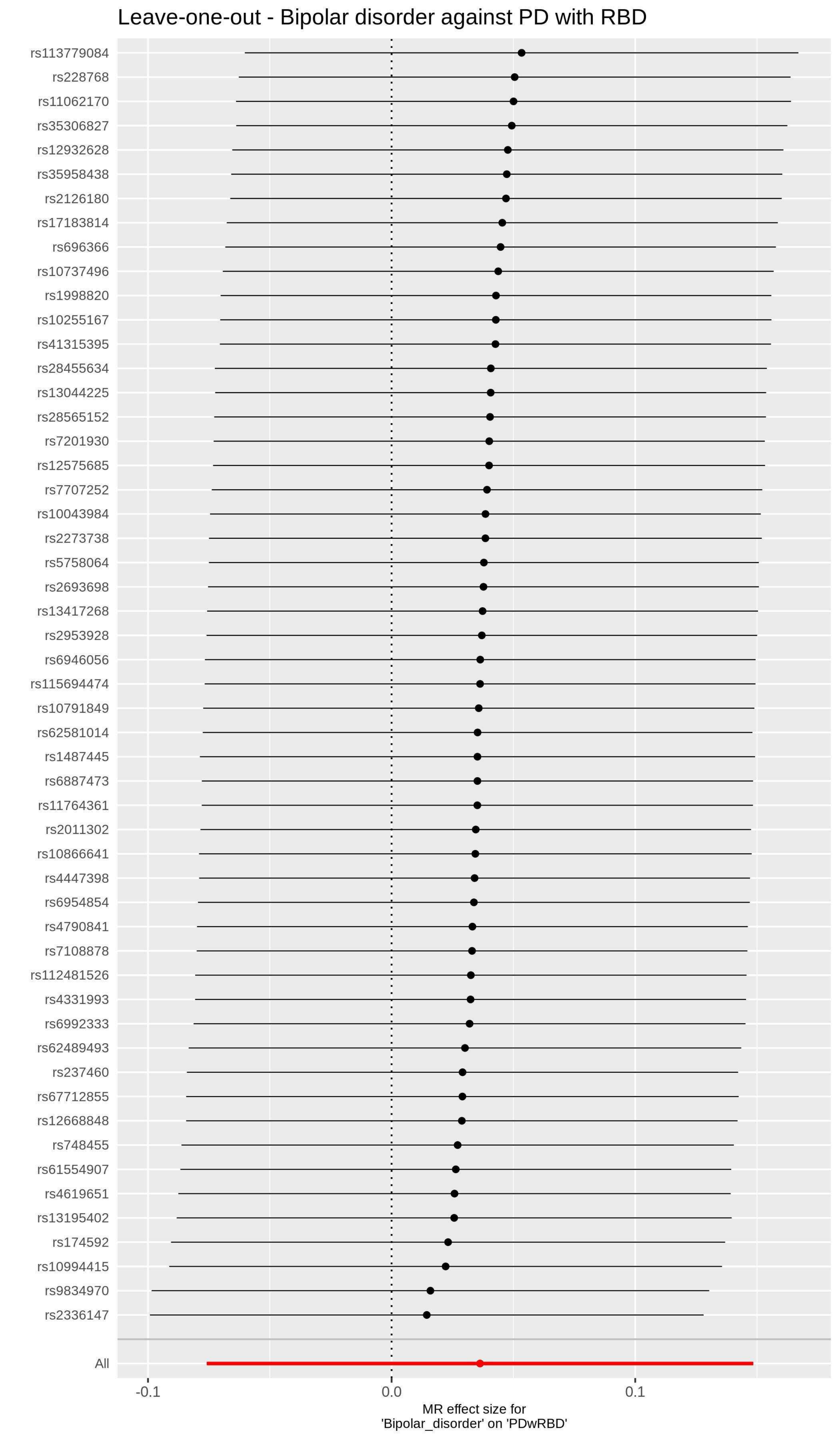

Leave-one-out - Bipolar disorder against PD with RBD
rs113779084
rs228768
rs11062170
rs35306827
rs12932628
rs35958438
rs2126180
rs17183814
rs696366
rs10737496
rs1998820
rs10255167
rs41315395
rs28455634
rs13044225
rs28565152
rs7201930
rs12575685
rs7707252
rs10043984
rs2273738
rs5758064
rs2693698
rs13417268
rs2953928
rs6946056
rs115694474
rs10791849
rs62581014
rs1487445
rs6887473
rs11764361
rs2011302
rs10866641
rs4447398
rs6954854
rs4790841
rs7108878
rs112481526
rs4331993
rs6992333
rs62489493
rs237460
rs67712855
rs12668848
rs748455
rs61554907
rs4619651
rs13195402
rs174592
rs10994415
rs9834970
rs2336147
All
-0.1
0.0
0.1
MR effect size for
'Bipolar_disorder' on 'PDwRBD'
